# Subcellular spatial transcriptomics reveals immune–stromal crosstalk within the synovium of patients with juvenile idiopathic arthritis

**DOI:** 10.1101/2025.08.05.25332835

**Authors:** Jun Inamo, Roselyn Fierkens, Michael R CLay, Anna Helena Jonsson, Clara Lin, Kari Hayes, Nathan Rogers, Heather Leach, Kentaro Yomogida

**Author notes:** These authors are co-corresponding authors (Jun Inamo, Kentaro Yomogida). Correspondence: Jun Inamo and Kentaro Yomogida.

## Abstract

Juvenile idiopathic arthritis (JIA) is the most prevalent chronic inflammatory arthritis of childhood, yet the spatial organization in the synovium remains poorly understood. Here, we perform subcellular-resolution spatial transcriptomic profiling of synovial tissue from patients with active JIA. We identify diverse immune and stromal cell populations and reconstruct spatially defined cellular niches. Applying a newly developed spatial colocalization analysis pipeline, we uncover microanatomical structures, including endothelial–fibroblast interactions mediated by NOTCH signalling, and a CXCL9–CXCR3 signaling axis between inflammatory macrophages and CD8+ T cells, alongside the characterization of other resident macrophage subsets. We also detect and characterize tertiary lymphoid structures marked by CXCL13–CXCR5 and CCL19-mediated signaling from Tph cells and immunoregulatory dendritic cells, analogous to those observed in other autoimmune diseases. Finally, comparative analysis with rheumatoid arthritis reveals JIA-enriched cell states, including *NOTCH3+* and *CXCL12+* sublining fibroblasts, suggesting potentially differential inflammatory programs in pediatric versus adult arthritis. These findings provide a spatially resolved molecular framework of JIA synovitis and introduce a generalizable computational pipeline for spatial colocalization analysis in tissue inflammation.

**One Sentence Summary:** Spatial transcriptomics reveals immune–stromal niches and disease-specific interactions in juvenile idiopathic arthritis synovial tissue.

## INTRODUCTION

Juvenile idiopathic arthritis (JIA) is the most common chronic arthritis of childhood, affecting 1.6-23 cases per 100,000 children(1, 2). It comprises a heterogeneous group of conditions classified into eight subtypes(3), of which oligoarticular and rheumatoid factor (RF)-negative polyarticular JIA (hereafter referred to as oligo/poly JIA) account for 60–80% of cases in North America(4). Recent advances in therapy have transformed the overall prognosis of JIA; however, a substantial proportion of children with oligo/poly JIA continue to experience chronic, relapsing disease courses(5, 6). The biological processes that determine the prognosis remain largely unknown.

Synovial tissue, a connective tissue lining the joints, is the primary site of the inflammation in JIA. However, previous studies on JIA have relied on synovial fluid due to practical difficulties of obtaining tissue. A recent landmark study provided the first cellular atlas of treatment-naive JIA synovium, combining Cellular indexing of transcriptomes and epitopes (CITE-seq) with *in situ* spatial transcriptomics platform that profiled 377 genes(7). This study highlighted the age-dependent differences in synovial cell populations and identified macrophage subsets associated with disease progression.

To deepen these insights, we sought to generate a comprehensive spatial and transcriptional profile of the JIA synovium. In this study, we applied a high-plex in situ transcriptomic platform (10x Genomics Xenium Prime 5K) to JIA synovial tissue, enabling the transcriptomes of thousands of cells to be measured within their native spatial organization at near single-cell (sub-10 μm) resolution in formalin-fixed, paraffin-embedded (FFPE) tissues. We developed an analytic pipeline tailored for high-resolution spatial transcriptome data, performing spatial neighborhood enrichment analyses at both cell-type and single-spot levels. Using this approach, we rediscovered the spatial interaction between fibroblasts and endothelial cells— previously identified in RA synovium(8)—in JIA tissues. Furthermore, we characterized macrophage and T-cell cross-talk and defined tertiary lymphoid structures (TLS) within the JIA synovial microenvironment, providing new insights into immune-stromal interactions in JIA.

## RESULTS

### High-resolution spatial profiling uncovers cellular architecture and disease-linked stromal population in JIA synovium

To explore the cellular diversity and spatial organization within the inflamed synovium of JIA patients, we employed advanced spatial transcriptomic profiling on synovial biopsy samples from nine patients with oligo/poly JIA, defined by International League of Associations for Rheumatology (ILAR) classification(3) (**Supplementary Table 1**). All oligoarticular JIA patients were classified as a persistent subtype at the time of biopsy. We collected synovial biopsies from treatment-naive patients (n=3) and from patients who had received various treatments, including intra-articular steroid injections, methotrexate, leflunomide and anti-TNF inhibitors (n=6). All patients were negative for rheumatoid factor. Six patients have normal C-reactive protein (CRP) levels, a finding consistent with previous reports indicating that 50-70% of patients with oligo/poly JIA exhibit normal CRP(9, 10). At the time of biopsy, all of our JIA samples exhibited active synovitis, as diagnosed by board-certified pediatric rheumatologists and supported by elevated Krenn inflammatory infiltrate scores (**Supplementary Table 1**).

Utilizing the 10X Xenium Prime 5K platform, we generated high-resolution spatial transcriptomic data capable of identifying discrete cell populations and characterizing their spatial interactions and niche formations within the tissue microenvironment (**Fig. 1a-c**). After preprocessing and batch correction steps (**Supplemental Figure 1a-c, Methods**), we performed unsupervised clustering of the spatial transcriptomic data (380,998 cells in total) and, based on manual annotation using canonical markers, delineated four major cellular compartments: T cell-innate lymphoid cells (ILCs), B/plasma cells, myeloid cells, and tissue-associated stromal cells, which included diverse subsets of fibroblasts and endothelial cells (**Fig. 1d-g**). Marker genes for each subcluster are listed in **Supplementary Table 2**.

**Fig. 1:**
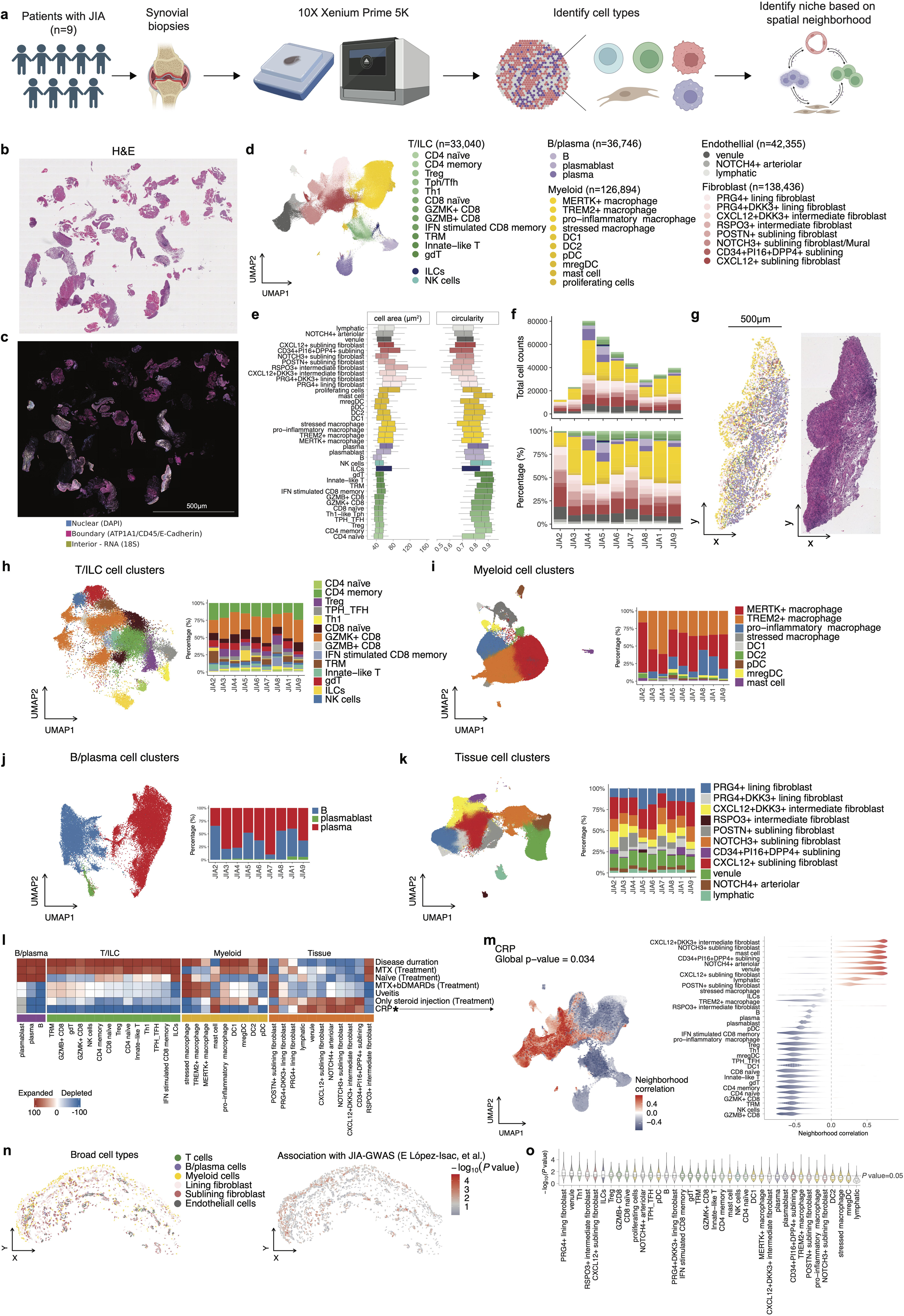
Spatial transcriptomic profiling of synovial tissue in JIA patients. **a**, Synovial biopsy samples were collected from nine patients diagnosed with JIA. Subcellular resolution spatial transcriptomic data was acquired from FFPE biopsy samples using the 10X Xenium Prime 5K platform. Cell types within synovial tissues were identified, and spatial neighborhoods were characterized based on spatial proximity analysis. **b-c**, Representative histological and spatial transcriptomic visualization of JIA synovium. Hematoxylin and eosin (H&E) staining of a representative synovial tissue sample (JIA4) (**b**), and corresponding spatial transcriptomic image visualized using Xenium Explorer (**c**). In panel **c**, Nuclear signals (DAPI) are shown in blue, cell boundary markers (ATP1A1, CD45, and E-cadherin) in magenta, and RNA signal (18S rRNA) in yellow. **d,** Identified major immune and tissue-associated cellular compartments in synovium, including T cells, B cells, myeloid cells, and stromal tissue cells (endothelial and fibroblast subsets). Within each broad cell type, fine-scale cell subpopulations were annotated. **e,** Box plots show the distribution of cell area (left) and circularity (right) across annotated cell populations. Circularity was calculated as (4 × π × Area) / (Perimeter^2^), with values closer to 1 indicating rounder cells. Box plots showing the median, interquartile range, and 1.5× interquartile range (IQR) whiskers. **f,** Composition of cell-types across samples. **g**, Representative example of the FFPE histological slide of synovial biopsy sample and spatial mapping of identified cell clusters. Colors are corresponding with panel **d**. **h-k,** Identified fine-scaled cell clusters on UMAP (left) and composition across samples (right) for T cells (**h**), myeloid cells (**i**), B/plasma cells (**j**), and stromal tissue cells (**k**). **l,** Heatmap of covarying neighborhood analysis (CNA) associations of specific cell states with each clinical variable. All testing was adjusted for age and sex. Colours represent the percentage of cell neighbourhoods from each cell state with positive phenotype correlations from white to red (expanded) or blue (depleted). Clinical variables associated globally (permutation p-value < 0.05) are annotated by asterisk (*). **m**, CNA for CRP association with cell states. Cells in UMAP are colored for expansion (red) or depletion (blue). **n**, Representative region, with cells colored by broad cell types (left) and the significance of the association (−log₁₀(P value)) with JIA derived from gsMap analysis (right). **o**, Violin plot showing the distribution of disease associations for each spatial spot across annotated synovial cell type, ordered from left to right by decreasing median association value.

T cell-ILC subpopulations included fine-scaled T cell subsets as well as ILCs (**Supplemental Figure 1d**). The CD8^+^ T cell population encompasses *CCR7*^+^ *TCF7* naïve CD8^+^ T cells, along with granzyme B-positive (*GZMB*^+^) and granzyme K-positive (*GZMK*^+^) CD8^+^ T cells, which have been previously characterized in the synovium of patients with RA(11). We also identified a distinct CD8^+^ T cell population, tissue-resident memory T cells (TRM), marked by expression of *ZNF683*, which encodes HOBIT(12). IFN stimulated CD8^+^ memory T cells were enriched with interferon stimulated genes such as *IFIT1* and *IFIT3*. CD4^+^ T cells consisted of *CCR7*^+^ *TCF7*^+^ naive CD4^+^ T cells, *FOXP3*^+^ regulatory T cells (Tregs)(13), and an *IFNG*^+^*TBX21*^+^ population designated as type 1 helper T cell (Th1). We also identified CD4^+^ T cells expressing *CXCL13* and *IL21*, corresponding to peripheral T cells and follicular T cells (T_PH_/T_FH_)(14). We further detected *TRDC*^+^ gamma-delta (γδ) T cells, and innate-like T cells expressing *ZBTB16* (encoding PLZF) and natural killer (NK) receptor *KLRB1*, with transcriptional profile resembling invariant natural killer T cells (iNKT cells)(15) and mucosal associated innate T (MAIT) cells(16). Innate subpopulations, NK cells and ILCs were also identified. ILCs were characterized by *NCAM1-*negative*, CD3E-*negative, and enrichment for *IL5* and *IL17A*, suggesting the presence of ILC2 and ILC3 subset(17).

B cell-plasma cells comprised *MS4A1*^+^ (encoding CD20) B cells, *CD38*^+^ plasma cells and *MKI67*^+^ plasmablast populations (**Supplemental Figure 1e**).

Myeloid cells encompassed diverse macrophage populations, dendritic cells (DCs), plasmacytoid dendritic cells (pDC), and mast cells (**Supplemental Figure 1f**). The macrophages clusters comprised several distinct clusters: *MERTK*^+^ macrophages, characterized by the enrichment of scavenger receptor *CD163* and *MRC1* (encoding CD206) and *MERTK*(*18*); *TREM2*^+^ macrophages, defined by high expression of *TREM2* as well as tissue-remodeling chemokines such as *MMP9*(*19*); pro-inflammatory macrophages expressing mediators such as *CXCL9*, *CXCL10* and *IL1B*, along with *CCR2*, a chemokine receptor indicative of infiltrating monocyte-derived macrophages(20–22). We also identified a subset of stressed macrophages marked by high expression of endoplasmic reticulum stress-related genes (*HYOU1*, *PDIA4*, *DDIT3*)(23). The DC cluster included *CLEC9A*^+^*XCR1*^+^*THBD*^+^ DC1, *CD1C*^+^*CLEC10A*^+^ DC2(24). Additionally, one DC cluster was characterized with *LAMP3*^+^, *CCR7*^+^ and immunomodulatory molecules such as *CD274* (encoding PD-L1), corresponding to recently described mature dendritic cells enriched in regulatory molecules (mregDCs)(25).

Stromal tissue-associated cell types included lining and sublining fibroblasts, and endothelial cells (**Supplemental Figure 1g**). FIbroblasts were classified into *PRG4*^+^ lining, *THY1*^+^ sublining fibroblasts and a mixture of *NOTCH3*^+^ sublining fibroblast and *MCAM*^+^ (encoding CD146) mural cells. Lining fibroblasts were further divided based on the expression of *DKK3*, which could play role in repair pathways(26), into *DKK3^low^*and *DKK3^high^ PRG4*^+^ subsets; and intermediate populations, which exhibited gene expression patterns between lining-sublining fibroblasts(27, 28), including *RSPO3*^+^, which could modulate Wnt/β-catenin signaling and have crucial effects on angiogenesis(29, 30). Sublining fibroblasts were further categorized based on the expression of *CXCL12*, *CD34, DKK3* and *POSTN*. Endothelial cells were classified into *PROX1*^+^*FLT4*^+^ lymphatic and blood vascular vessels, the latter were further divided into *LIFR*^+^ *SELE*^+^ venules and *NOTCH4*^+^ arteriolar endothelial cells.

To investigate the cellular sources of cytokines and receptors relevant to synovial inflammation, we examined the expression of genes from a curated cytokine signaling gene set(31) (**Supplemental Figure 1h**). Most cytokines and receptors showed dominant expression within specific cell populations; for instance, fibroblasts and B cells predominated among cells detectable *IL6*, whereas *IL18* expression was primarily restricted to macrophages and dendritic cells.

To quantitatively characterize morphological differences across cell types, we computed cell area and circularity (**Fig. 1e**). Stromal cells such as fibroblasts and endothelial subsets exhibited larger and more variable cell areas compared to immune cells, whereas lymphocyte populations demonstrated higher circularity consistent with their small, rounded morphology.

Quantitative assessment across individual biopsy samples revealed variability in cellular composition, with significant inter-patient heterogeneity observed in the proportions of immune and stromal cells (**Fig. 1h-k**). Patients with higher inflammation state defined by serum CRP levels exhibited enrichment of sublining/intermediate fibroblast and endothelial cell (venules and arteriolar endothelium) populations relative to total synovial cells, indicating a potential critical role for these cells in potentiating inflammation in JIA synovium (**Fig. 1l-m**), as sublining fibroblasts are known as a resource of inflammatory molecules in RA-synovium(28, 32).

To investigate the spatial genetic relevance of cell populations within the synovium, we applied gsMap(33), a recently developed statistical framework that integrates spatial transcriptomic data with GWAS through stratified linkage disequilibrium (LD) score regression (sLDSC). This approach enables the mapping of disease heritability enrichment onto specific spatially resolved cell populations based on GWAS summary statistics of JIA(34). Notably, stromal populations—not only intermediate/sublining fibroblasts and vascular endothelial subsets, but also lining fibroblast and T cell subsets—exhibited significant enrichment of JIA-associated genetic signals (**Fig. 1n-o**). This suggests that these spatially localized niches, in particular stromal cells, may contribute to not only inflammatory activity but also disease susceptibility via genetically driven mechanisms.

Together, these results offer a detailed characterization of the spatially resolved cellular landscape of JIA synovium and their association with clinical indicators and disease-susceptibility.

### Spatially resolved niches reveal distinct immune–stromal compositions and gene programs

To identify tissue microenvironments with distinct immune and stromal compositions, we defined spatial “niches” based on local co-occurrence patterns of cell clusters within the synovium (**Methods**). A spatial “niche” was defined here for each cell by examining the composition of its 30 nearest spatial neighbors, summarizing the local abundance of each cell type. Cells with similar neighborhood compositions were grouped using k-means clustering to delineate discrete spatial niches. Each niche was then annotated by its relative enrichment in major cell lineages, resulting in four biologically interpretable niche classes: T+B/plasma cell niche, Myeloid+Stromal cell niche, Myeloid+T cell niche, and Stromal niche (**Fig. 2a-b, Supplemental Figure 2a**). Composition of annotated niches showed variability across patients with JIA (**Supplemental Figure 2b**). Analysis of these niche classes revealed distinct associations with patient clinical features (**Fig. 2a**). However, significant cellular and functional heterogeneity was observed even within these major classes. For example, the Myeloid+Stromal cell niche contained functionally diverse sub-niches: some were enriched in macrophages with high systemic inflammation (e.g., Niche 10), while others were dominated by PRG4+ lining fibroblasts and exhibited a less inflammatory profile (e.g., Niche 22). This intra-niche heterogeneity highlights a granular spatial organization where specialized microenvironments exist within a single tissue compartment.

**Fig. 2:**
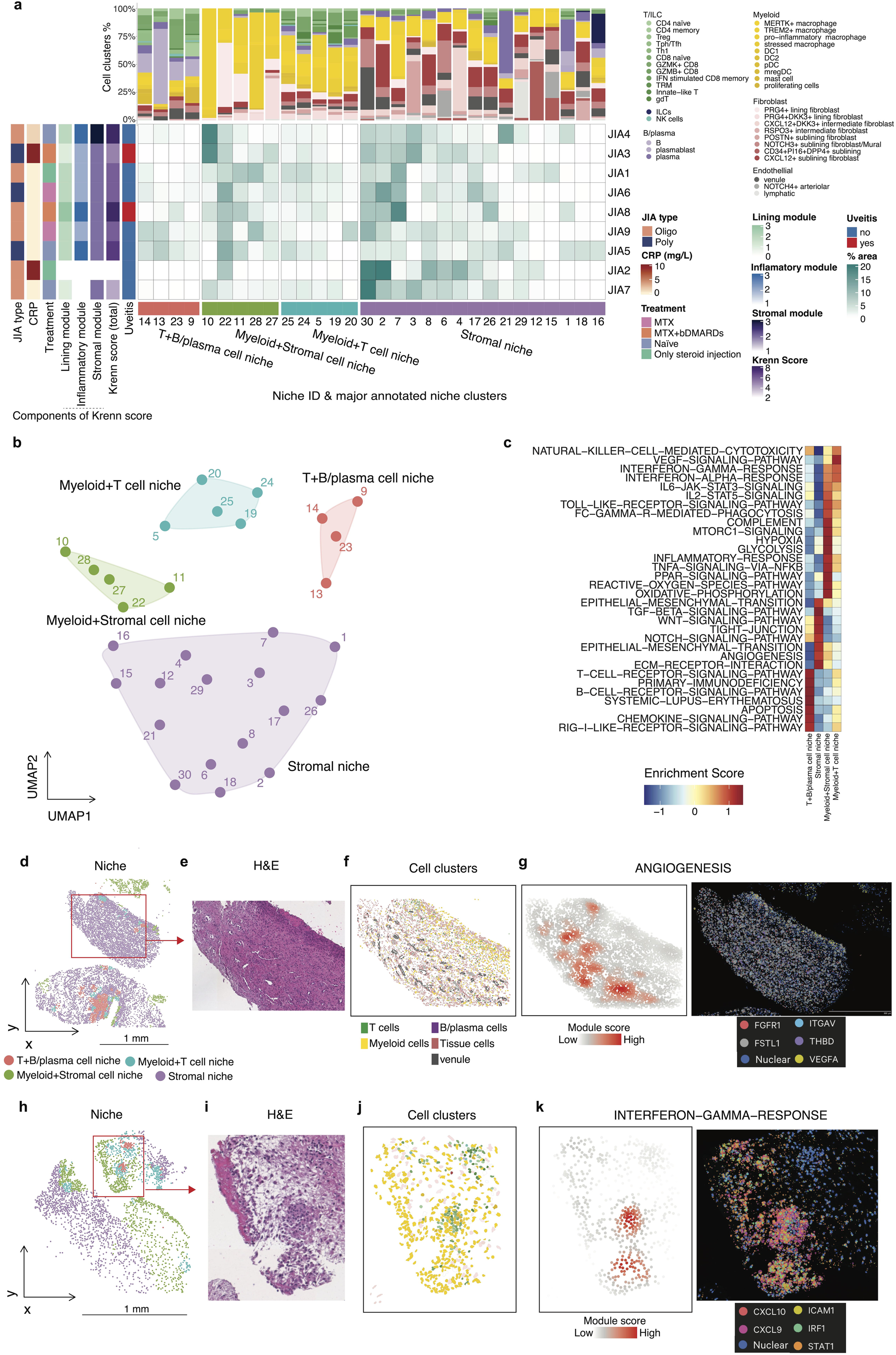
Characterization of spatial niches and their associated pathway activities in JIA synovium. **a**, The central heatmap displays the percentage area of 30 spatial niches (columns) for each patient (rows). Patient samples are annotated by clinical features on the left. The bar plots above illustrate the proportional composition of cell clusters for each corresponding niche. Spatial niches are grouped by the major annotated niche clusters shown at the bottom. **b,** UMAP embedding of niches based on their immune-stromal cell composition. Each dot represents a niche and is colored by an annotated niche cluster. **c,** Heatmap showing normalized gene set enrichment scores for related pathways across the four annotated niche classes. **d–e,** Representative images of association between niche clusters and ANGIOGENESIS pathway. Spatial mapping of annotated niches (**d**) from a representative synovial tissue section and H&E staining (**e**). **f**, Spatial distribution of cell clusters in the zoom-in of the section in panel **d**. **g**, Left, spatial enrichment score of the ANGIOGENESIS pathway. Right, Corresponding DAPI and expression of representative genes of the pathway. **h–i,** Representative images of association between niche clusters and INTERFERON−GAMMA−RESPONSE pathway. Spatial mapping of annotated niches (**h**) from a representative synovial tissue section and H&E staining (**i**). **j**, Spatial distribution of cell clusters in the zoom-in of the section in panel **h**. **k**, Left, spatial enrichment score of the INTERFERON−GAMMA−RESPONSE pathway. Right, Corresponding DAPI and expression of representative genes of the pathway.

To investigate the functional states of these niches, we performed pathway enrichment analysis comparing niche clusters. This analysis revealed class-dominant pathway signatures, including IFN-α/γ signaling in Myeloid-T cell–enriched niches, inflammatory and phagocytosis-related signatures in Myeloid-Stromal-rich niches, T-and B-cell-receptor signalling in B-T cell-enriched niches, and angiogenesis-related pathways in stromal niches (**Fig. 2c**). In addition, several other signaling cascades known to be implicated in JIA pathogenesis were selectively enriched across spatial niches. For instance, the IL6–JAK–STAT3 pathways—critical in T cell differentiation(35) and chronic inflammation(36)— as well as complement components, which have been previously associated with the disease marker in JIA(37), were elevated in Myeloid–Stromal cell niches. Meanwhile, TGF-β and WNT signaling pathways, implicated in fibroblast activation and tissue fibrosis(38, 39), and fibroblast-mediated inflammation and treatment-refractory in RA(40, 41), were more pronounced in stromal niches.

Histological mapping of niches and individual cell clusters demonstrated their anatomical relevance. For instance, stromal niches exhibited localized enrichment of the angiogenesis pathway, as visualized by spatial gene set scores (**Fig. 2d-g**). Another histological section also showed enrichment of IFN-γ signaling in Myeloid-T and Myeloid-Stromal-rich niches (**Fig. 2h-k**). These findings highlight the spatial heterogeneity of the synovial immune microenvironment in JIA and demonstrate that spatial niches are associated with distinct cell-type compositions and molecular programs.

### Spatial neighborhood enrichment analysis reveals NOTCH3-mediated interactions between endothelial cells and sublining fibroblasts in JIA synovium analogous to RA

Since tissue-microenvironment shapes both the cell identity and localization, we aimed to characterize the cell-cell interaction in each niche based on spatial proximity. To achieve this, we developed a custom pipeline to statistically test the spatial proximity between these cell types (**Fig. 3a**, **Methods**). We tested the performance of the pipeline using simulation data, assuming proximity pattern as concentric circle and layer, and found that it could efficiently identify pairs of cell types that were close to each other within 30 μm (**Fig. 3b-c**). To validate our spatial neighborhood analysis pipeline, we further utilized publicly available spatial transcriptomic data of human breast cancer and mouse brain (**Supplemental Figure 3a-d**). Spatial neighborhood analysis revealed significant enrichment in proximity such as between cancer-associated fibroblasts (CAFs) and cytotoxic T cells and astrocytes and oligodendrocytes, respectively, underscoring their spatial interplay in the tissue architecture as described in the published study(42, 43). The human breast cancer and mouse brain datasets comprised 8,273 and 17,215 cells, respectively, and the analysis, using four parallel threads, was completed in 7.6 seconds and 31.7 seconds, respectively, demonstrating the computational efficiency of the pipeline.

**Fig. 3:**
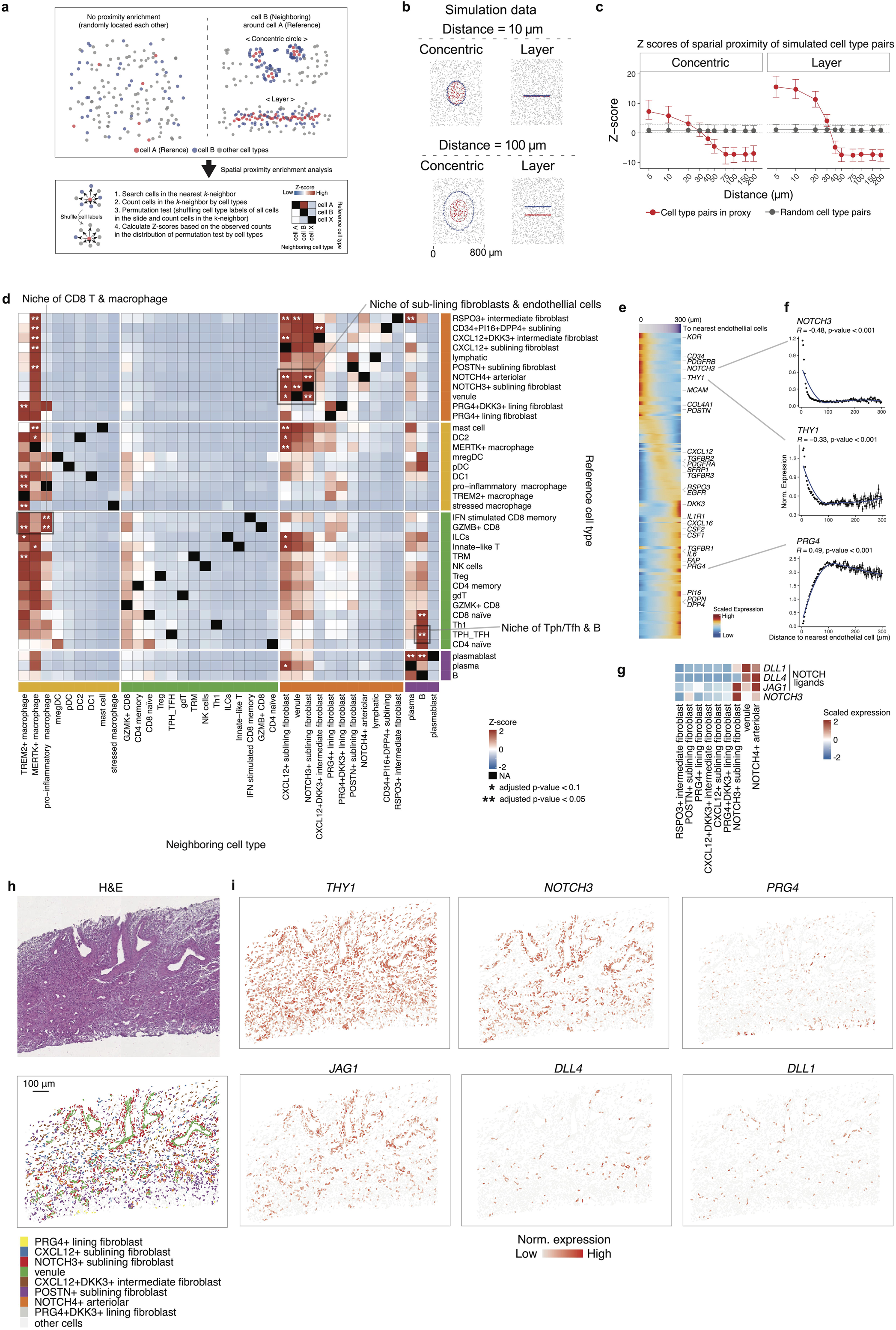
Spatial neighborhood enrichment and niche characterization of sublining fibroblasts and endothelial cells in JIA synovium. **a**, Schematic description of the spatial neighborhood enrichment analysis pipeline. The procedure includes identifying nearest neighbors, quantifying neighborhood cell-type composition, performing permutation tests by shuffling cell-type labels, and calculating enrichment Z-scores. **b**, Examples of two simulation patterns (concentric circles and layered arrangement) utilized to evaluate the performance of the spatial neighborhood enrichment method. **c**, Results from the simulation experiments. The x-axis represents pre-specified actual proximity distances, and the y-axis shows calculated spatial enrichment Z-scores. The red line indicates Z-scores for cell types that were designed to be spatially close, whereas the grey line represents randomly distributed cell types. The grey dashed line indicates the multiple testing-adjusted significance threshold for Z-scores. **d**, Spatial neighborhood enrichment using our data from samples of JIA synovium. Statistical significance is indicated as * (adjusted p-value < 0.1) and ** (adjusted p-value < 0.05). The highlighted grey square indicates the niche of sublining fibroblasts and endothelial cells. *P*-values were adjusted by the Benjamini-Hochberg method. **e**, Characterization of the identified niche of sublining fibroblasts and endothelial cells. The heatmap displays the relationship between distances to nearest endothelial cells (purple bar above) and fibroblast gene expression. Rows represent fibroblast-associated genes, and columns represent individual cells. Fibroblast marker genes are labeled. **f**. Line plots depicting binned distance to nearest endothelial cells (x-axis) and normalized gene expression (y-axis) of representative fibroblast genes. Statistical significance was assessed using Spearman correlation coefficients and p-values. **g**, Heatmap of gene expressions related to Notch signaling pathway components across different cell types. **h**, Representative spatial coordinates of an identified niche of sublining fibroblasts and endothelial cells, matching pathological images. Cells are colored according to their cluster assignments. **i**. Spatial expression patterns of fibroblast-related and Notch signaling-related genes within the identified niche location.

We applied this spatial neighborhood enrichment analysis pipeline to our JIA synovium spatial transcriptomic data and identified significant spatial proximities such as those between endothelial cells and sublining fibroblasts (**Fig. 3d**). Given previous findings that endothelial cells and fibroblasts interact via NOTCH3 signaling in RA synovium(8), we further investigated whether a similar interaction exists in JIA. Indeed, we observed that *THY1*^+^ sublining fibroblasts were enriched in proximity to endothelial cells and simultaneously exhibited elevated *NOTCH3* expression (**Fig. 3e-f**). Consistently, endothelial cells highly expressed the NOTCH3 ligands, such as *DLL1/4* and *JAG1* (**Fig. 3g**). Representative regions highlighting the spatial niche established by these interacting cell populations demonstrated the co-localization and reciprocal signaling between endothelial cells and sublining fibroblasts (**Fig. 3h-i, Supplemental Figure 3e-f**). This observation suggests an organized microanatomical structure within the synovial tissue, reinforcing the notion that endothelial–fibroblast interactions mediated through NOTCH3 signaling might contribute to the formation and maintenance of inflammatory niches in JIA synovium.

### Spatial crosstalk with T cells and vasculature orchestrates macrophage polarization

Given the established importance of macrophages in the pathogenesis of inflammatory arthritis, we aimed to explore how macrophage gene expression is altered in relation to their spatial proximity to other cell types within the synovial tissues. Since monocytes enter through vasculature and differentiate to synovial macrophages(44–46), we first examined how gene expressions in macrophages correlated with their distance to endothelial cells (**Fig. 4a**). Anti-inflammatory or tissue-repairing markers such as *MRC1*, *CSF1R*, and *MERTK*, which were known to be expressed on healthy synovial tissue macrophages(47) and increased in synovium of RA patients with remission states(18), showed higher expression in macrophages proximal to vasculature, whereas expression of pro-inflammatory-associated genes including *IL1RN*, *NFKB2*, and *JAK2*, increased with distance from the nearest endothelial cell. We also noted that *TREM2*, a gene traditionally associated with tissue-repairing macrophage programs(48), exhibited a distinct spatial pattern compared to other anti-inflammatory markers, as *TREM2* expression increased with distance from endothelial cells. *CD14* expression decreased while *CD68* expression concurrently increased with distance from endothelial cells, suggesting that circulating monocytes progressively differentiate into macrophages after entry through the endothelium as they adapt to the synovial microenvironment(44–46).

**Fig. 4:**
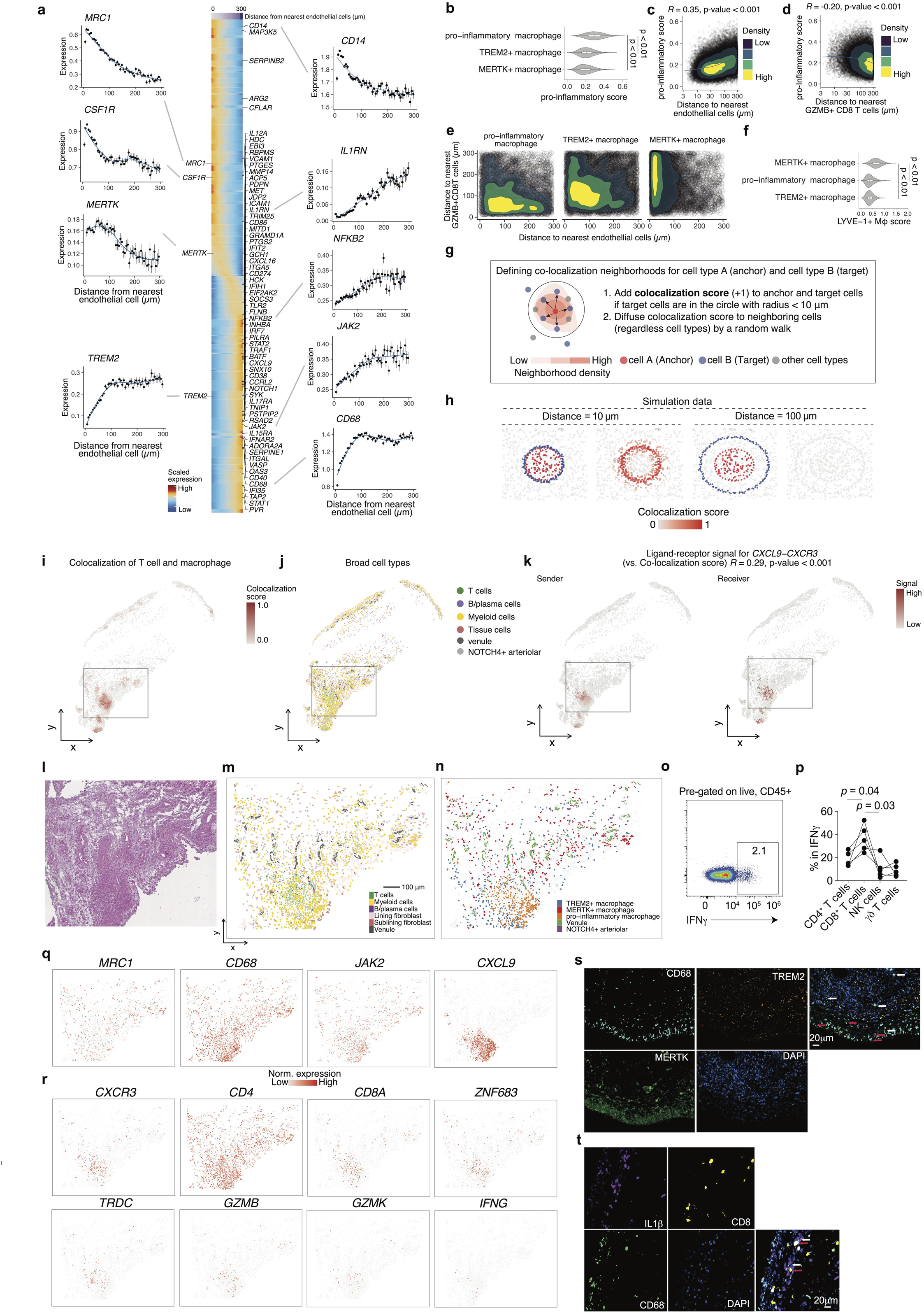
Spatial distribution and interactions of distinct macrophage states in JIA synovium. **a**, Scaled gene expression of selected macrophage markers plotted as a function of distance from the nearest endothelial cell. Center heatmap shows scaled expression of macrophage-related genes in individual cells ordered by proximity to endothelial cells (0–300 µm), with representative genes annotated. **b**, Violin plots showing the distribution of pro-inflammatory module scores across macrophage subtypes. *P* values were determined by the Wilcoxon rank-sum test. **c–d**, Correlation between pro-inflammatory module scores of individual macrophage cells (each point) (y-axis) and distance to the endothelial cells (**c**) or *GZMB*^+^ CD8 T cells (**d**) (x-axis). Color-filled contours represent two-dimensional kernel density estimates. Spearman’s correlation coefficient and p-values are shown. **e**, Density plots depict the distances of individual macrophage cells (each point) to the nearest endothelial cells (x-axis) and *GZMB*^+^ CD8 T cells (y-axis) by macrophage subtypes. Color-filled contours represent two-dimensional kernel density estimates. **f**, Violin plots showing the distribution of LYVE-1^+^ macrophage module scores across macrophage subtypes. *P* values were determined by the Wilcoxon rank-sum test. **g**, Schematic illustrating spatial colocalization scoring between anchor cells (e.g., macrophages) and target cells (e.g., T cells) within a 10 µm radius. **h**, Representative spatial maps of high-and low-co-localization scores between anchor and target cells across tissue sections by simulated patterns. **i**, Spatial heatmap of colocalization scores between T cells and macrophages. **j**, Spatial map of broad cell type annotation including T cells, myeloid cells, B/plasma cells, and tissue cells. **k**, Spatial ligand– receptor analysis results for CXCL9–CXCR3, which exhibited the strongest correlation with macrophage–T cell co-localization scores. **l-n**, Zoom-in of the highlighted grey square region in panel **i-k** showing identified niche of T cells, macrophages, and endothelial cells. H&E staining image (**l**) and spatial mapping colored according to their cluster assignments (**m-n**). **o**, **o-p**, Representative flow cytometry plot showing intracellular IFNγ expression (**o**) and quantification of the frequencies of CD4^+^ T cells, CD8^+^ T cells, NK cells, and γδ T cells among IFNγ^+^ cells (**p**) from the synovial fluid of oligo/poly JIA patients (n = 4). Each line connects measurements from the same individual across different cell types. *P* values were determined by Friedman’s test. **q–r**, Same region as in panel **l-n**, spatial expression of representative marker genes for macrophages (**q**) and T cells (**r**). Color scale indicates normalized expression levels. **s**, Representative immunofluorescence image showing the localization of CD68^+^MERTK*^+^* macrophages (white arrows) and CD68^+^TREM2*^+^* macrophages (red arrows) in the synovial tissue. Nuclei were counterstained with DAPI (blue). **t**, Representative immunofluorescence image demonstrating physical colocalization of CD8^+^ T cells (red arrows) and IL1β^+^CD68^+^ pro-inflammatory macrophages (white arrow) within inflammatory niches of the synovium. Nuclei were counterstained with DAPI (blue). Images are representative of n = 5 independent samples.

Next, we aimed to characterize the microenvironmental cues that induce inflammatory gene expression in macrophages. As expected from our definitions of cell type clusters, pro-inflammatory macrophages exhibited the highest pro-inflammatory module scores(49, 50) (**Fig. 4b**). We observed a positive correlation between pro-inflammatory polarization and distance from endothelial cells (*R* = 0.35, p < 0.001) (**Fig. 4c**). Since we found that pro-inflammatory macrophages are co-localized with *GZMB*^+^ CD8^+^ T cells (**Fig. 3d**), we asked whether inflammatory gene expression in macrophages is induced when they are in close proximity to CD8^+^ T cells. Indeed, pro-inflammatory scores in macrophages gradually decreased along with distance from *GZMB*^+^ CD8^+^ T cells (*R* = −0.20, p < 0.001) (**Fig. 4d**), and a similar trend was observed for *GZMK*^+^ CD8^+^ T cells (*R* = −0.11, p < 0.001) (**Supplemental Figure 4a**). Importantly, two-dimensional density contour plots revealed distinct spatial patterns across macrophage subsets (**Fig. 4e**); pro-inflammatory macrophages were located closer to *GZMB^+^*CD8^+^ T cells than endothelial cells.

In contrast, *MERTK*^+^ macrophages showed a strong spatial association with endothelial cells. To further investigate the potential identity of *MERTK*^+^ macrophages, we examined their similarity to LYVE-1^+^ perivascular macrophages, which have been previously described as collagen-remodeling cells positioned near blood vessels in steady-state tissues. As LYVE-1 itself was not included in our Xenium probe panel, we inferred LYVE-1^+^–like phenotype by computing gene module scores across macrophage clusters using a gene signature associated with LYVE-1^+^ macrophages(18). The analysis revealed that *MERTK*^+^ macrophages exhibited the highest LYVE-1^+^ macrophage score among the identified subsets (**Fig. 4f**). This result supports the notion that *MERTK*^+^ macrophages in JIA synovium represent a transcriptionally related population to homeostatic LYVE-1^+^ perivascular macrophages, potentially involved in vascular support and extracellular matrix turnover.

Given the reciprocal interactions of macrophages and fibroblasts(51), we examined their spatial relationships with lining fibroblasts, and observed that *TREM2^+^*macrophages were enriched in regions close to lining fibroblasts (<50 μm) (**Supplemental Figure 4b**). To further characterize *TREM2^+^* macrophage populations, we computed a module score using the CX3CR1^+^ lining macrophage gene signature previously defined in a murine model of joint inflammation(52). We observed significantly higher CX3CR1^+^ lining macrophage scores in *TREM2*^+^ macrophages, compared to other macrophage subsets (**Supplemental Figure 4c**). These results suggest that *TREM2*^+^ macrophages in our dataset may represent a human equivalent of CX3CR1^+^ barrier-forming macrophages, supporting their putative role in maintaining tissue homeostasis and limiting inflammatory responses at the synovial interface(52, 53).

Although our spatial neighborhood enrichment analysis effectively captured cell-type interactions, it did not resolve individual cell-level proximities. To overcome this limitation and enable identification of direct cellular interactions at single-spot resolution, we developed a colocalization score designed to precisely capture interactions between specific cell populations within spatial transcriptomics data (**Fig. 4g, Methods**). We implemented this new scoring method, leveraging spatial coordinates to assign higher scores to cells that coexist closely within the spatial neighborhood. Using simulated and public spatial transcriptomics data, we confirmed the robustness of this approach, demonstrating that our colocalization score accurately identified known pairs of spatially proximal cells under various controlled scenarios (**Fig. 4h, Supplemental Figure 4d-e**).

Subsequently, we utilized the colocalization score to investigate ligand–receptor signaling interactions between macrophages and T cells by integrating ligand–receptor pair predictions from the COMMOT pipeline(54) (**Supplemental Figure 4f**). The distribution of correlation *p*-values showed ligand-receptor interaction signals that were spatially covaried with macrophage-T cell colocalization scores (**Supplemental Figure 4g**). CXCL9-CXCR3 ligand-receptor pair exhibited the highest correlation with the macrophage-T cell colocalization scores, implicating this chemokine signaling axis as a potential critical mediator facilitating spatially organized crosstalk between these immune cells within JIA synovium (**Fig. 4i-k, Supplemental Figure 4h-j**).

We next focused on the niches composed primarily of macrophages, endothelial cells and T cells (**Fig. 4l-n, Supplemental Figure 4k**). While both *MERTK^+^* and *TREM2^+^*macrophages were distributed across the lining and sublining layers, these subsets exhibited partially distinct spatial patterns; *MERTK^+^* macrophages showed local enrichment around endothelial cells, whereas *TREM2^+^* macrophages were broadly present in both lining and sublining areas. This distribution of *TREM2^+^*macrophages is consistent with findings in RA, where they are observed in both compartments in the active disease state(18). On the other hand, pro-inflammatory macrophages were colocalized with T cells. Within the Myeloid-T cell niche, *GZMB*^+^ CD8^+^ T cells exhibited the high expression of *IFNG* transcripts, which drives macrophage polarization toward a pro-inflammatory phenotype (**Supplemental Figure 4l**). We further quantified IFNγ production by lymphocyte subsets in synovial fluid samples from oligo/poly JIA patients, and we confirmed that CD8^+^ T cells are the predominant source of IFNγ (**Fig. 4o-p**). Spatial transcriptomic mapping demonstrated enriched expression of macrophage-related inflammatory genes precisely at these niches (**Fig. 4q-r, Supplemental Figure 4m**). Immunofluorescence staining of active JIA synovial samples confirmed that *MERTK^+^* macrophages exhibited focal accumulation within the sublining layer, likely around endothelial cells, whereas *TREM2^+^* macrophages dispersed in both lining and sublining layers (**Fig. 4s**). Additionally, physical colocalization of CD8^+^ T cells with pro-inflammatory macrophages (**Fig. 4t**) was observed in JIA synovial tissue, providing orthogonal validation of these inflammatory niches in situ.

Taken together, these findings underscore the spatial diversity of macrophage subsets in JIA synovium, and suggest a dynamic phenotypical change in shaping the inflammatory macrophage phenotype via macrophage–T cell interactions, particularly through chemokine and cytokine signaling.

### Spatial identification and cellular characterization of tertiary lymphoid structures in JIA synovium using T and B cell colocalization scores

Given the established roles of TLS in chronic inflammatory conditions(55–58), we investigated their presence and cellular composition within JIA synovium. First, we applied the colocalization score framework to specifically assess interactions between T cells and B/plasma cells in order to detect potential TLS regions. These computationally identified regions matched with validated TLS regions by independent histopathological analysis performed by a blinded pathologist and T-B/plasma-rich regions by spatial mapping of broad cell types (**Fig. 5a-c**).

**Fig. 5:**
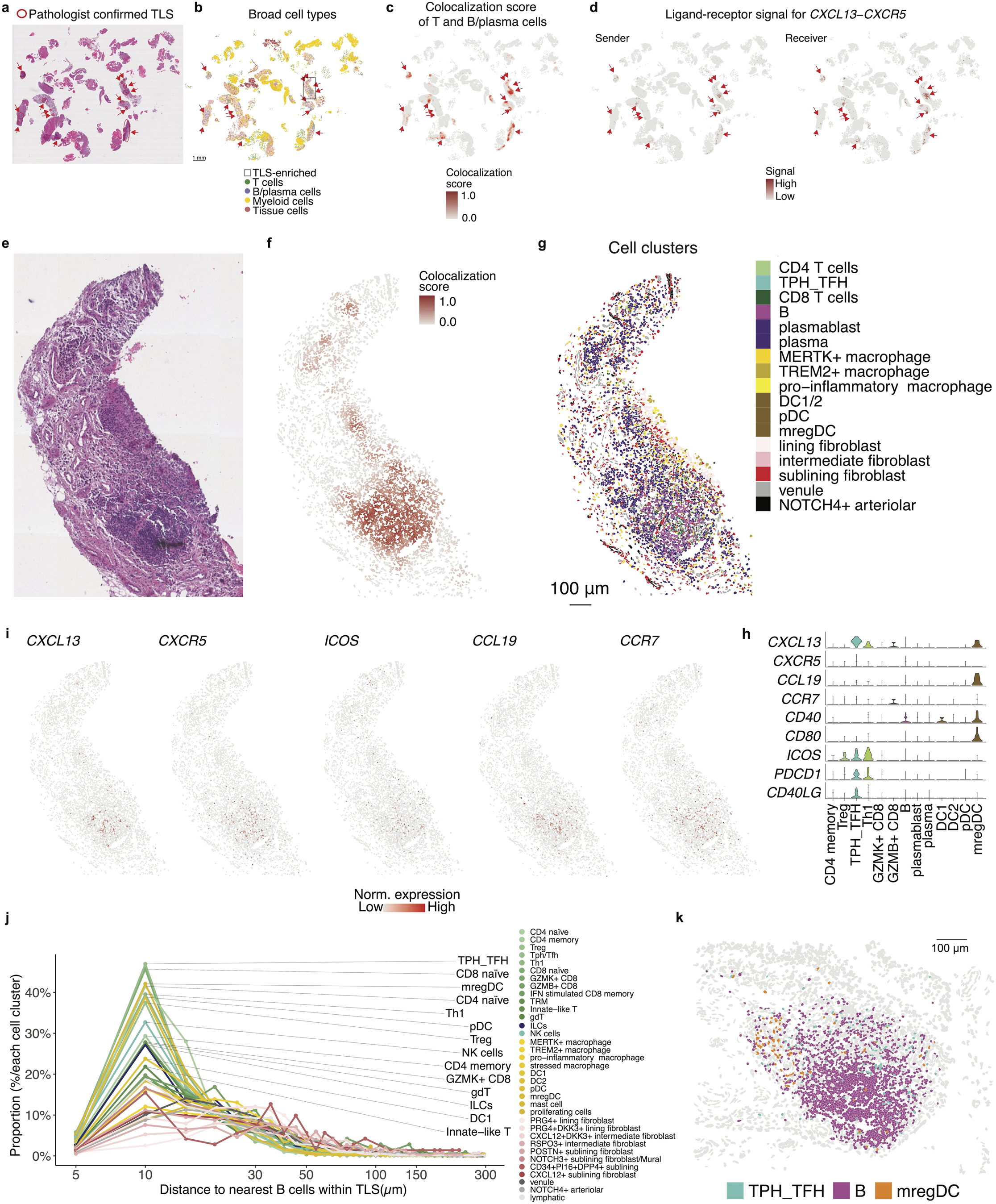
Identification and characterization of tertiary lymphoid structures (TLS) using T and B cell colocalization scores. **a**, H&E staining images of synovial tissue, illustrating representative TLS as identified by a blinded pathologist. **b**, Spatial distribution of T and B cells. Scale bar, 1 mm. The highlighted grey square indicates the TLS-enriched region. **c**, Colocalization scores for T and B cells. **d**, Spatial *CXCL13*-*CXCR5* interaction signals. **e**, H&E staining image of zoomed-in of the area with the highest TLS density highlighted in panel **b. f-g**, Same region as in panel **e**, colored by T and B cell colocalization score (**f**) and fine-scale cell cluster assignments (**g**). **h**, Violin plots showing the expression levels of key TLS-related genes across different cell clusters identified within the TLS. **i**, Spatial expression patterns of TLS-associated genes. **j**, Line plot showing the proportion of each immune cell cluster (y-axis, % of total cells within each cluster) relative to its binned distance to the nearest B cell within identified TLS regions (x-axis, log-scaled). Each line represents a distinct immune cluster, with colors corresponding to the cluster identities. Points denote the proportion per 5-μm distance bin, and annotations on the right indicate the top clusters based on peak proximity to B cells. **k**, Representative TLS region showing colocalization of Tph/Tfh cells, B cells, and mregDCs.

To assess the molecular mechanisms underlying TLS formation, we examined ligand– receptor signaling between T cells and B cells within identified TLS. The chemokine signaling axis CXCL13–CXCR5 showed significant enrichment in these TLS regions, suggesting a shared mechanism of TLS formation and maintenance with other chronic diseases (**Fig. 5d**).

Histological image (**Fig. 5e**), spatial visualization of TLS colocalization scores (**Fig. 5f, Supplemental Figure 5**) and distribution of cell clusters confirmed the enrichment of Tph/Tfh cells within TLS-dense regions (**Fig. 5g**). These clusters co-expressed high levels of key TLS-related genes including *CXCL13, CXCR5, ICOS,* and *CCR7* (**Fig. 5h-i**). Additionally, *CCL19* expression, ligand for *CCR7* and likely from mregDC, further supports the establishment of TLS-associated chemotactic gradients. The elevated expression of *CD40* and *CD80* in mregDCs may facilitate the priming of naive T cells.

To further characterize the spatial organization of cell types within TLS, we quantified the distance from each cell to its nearest B cell across identified TLS regions and calculated the distribution of cell types across distance. This analysis revealed that several immune cell populations, particularly T cells as expected, including Tph/Tfh cells as the most enriched cell types, proxy for B cells (**Fig. 5j**). Other than T cell clusters, mregDCs were highly enriched in close proximity (within ∼10 μm) to B cells (**Fig. 5k**). These spatial patterns suggest their functional involvement in TLS formation and maintenance, likely through CXCL13–CXCR5 (T-B) and CCL19-mediated signaling from mregDCs.

Together, these findings demonstrate the utility of the colocalization score in identifying TLS in JIA synovium and provide insights into the complex cellular interactions and signaling pathways driving TLS formation and maintenance.

### Integrated comparison of JIA and RA synovial tissue reveals potential disease-specific cellular niches

To compare the cellular architecture of JIA and RA synovium, we integrated our spatial transcriptomic data with publicly available RA synovial scRNA-seq data(28) (**Methods**). RA-derived cell clusters were transferred to JIA cells in the shared latent space (**Fig. 6a**). The transferred cell-type labels showed high concordance with our annotations for major immune and stromal compartments in JIA synovium, including T cells/ILCs, B/plasma cells, myeloid cells, stromal tissue cells (**Fig. 6b**), with detailed results for each lineage shown in **Supplemental Figure 6**.

**Fig. 6:**
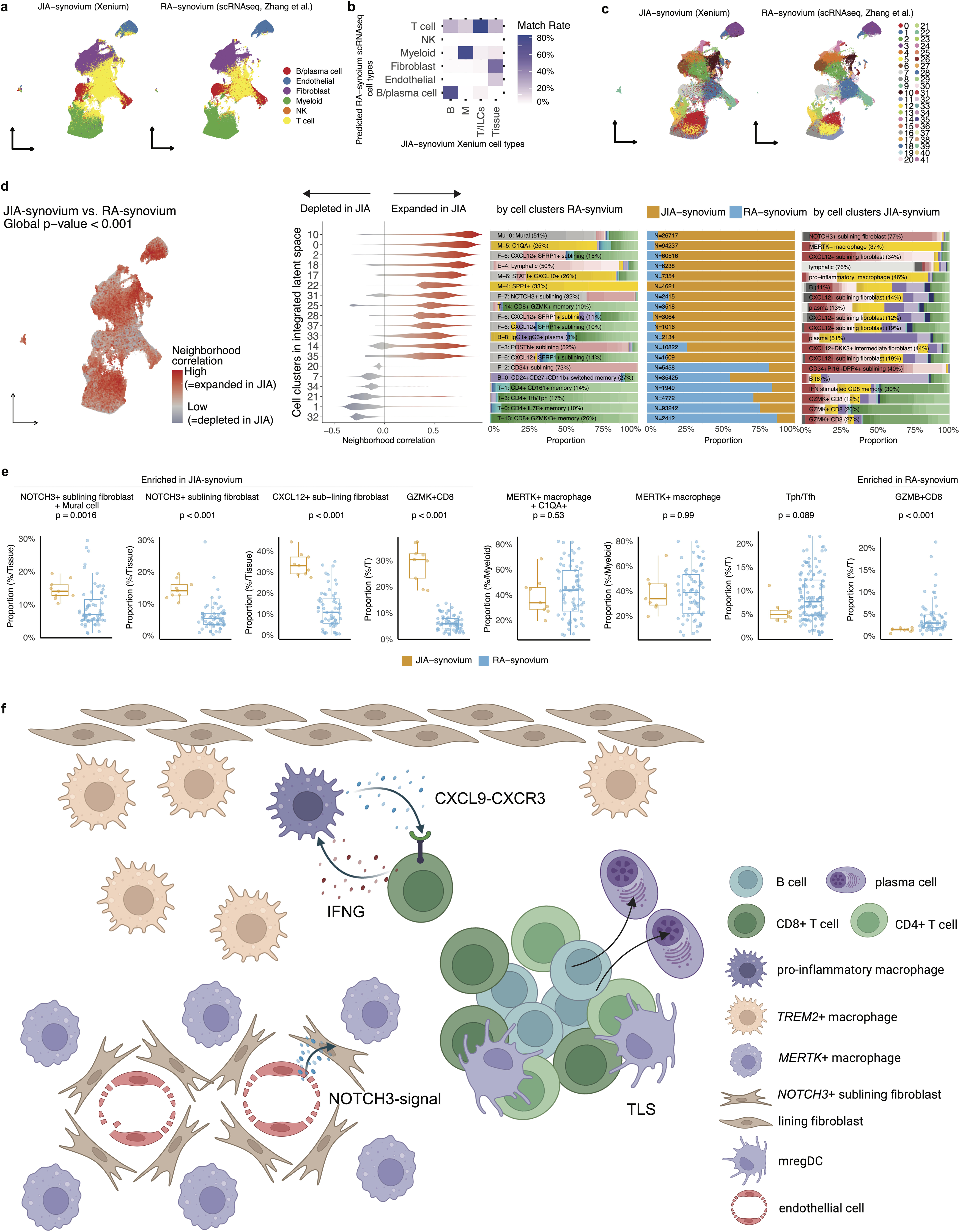
Comparative analysis of JIA and RA synovium using integrated spatial-transcriptomic and scRNA-seq data. **a**, Cell-type level label transfer results, where RA synovium scRNA-seq (Zhang et al.(28)) was used as the reference and JIA synovium Xenium data as the query. RA-derived annotations were transferred to individual JIA cells using a k-nearest neighbors (kNN) approach in the integrated latent space. **b**, Match rate between transferred RA-synovium cell types and our original JIA-synovium-based annotations. Matrix shows percentage overlap between each transferred and original broad cell type. **c**, Cell clustering in the integrated latent space. UMAP visualizations of both JIA (Xenium) and RA (scRNA-seq) cells are colored by newly defined shared clusters. **d**, CNA identifies integrated-space clusters that are expanded or depleted in JIA synovium relative to RA synovium. Sex and disease duration were adjusted for, while age was excluded from covariates due to its collinearity with disease status, reflecting the distinct age of onset between RA and JIA. Left: Spatial visualization of cells colored by neighborhood correlation (red = expanded in JIA, blue = depleted). Middle: Violin plots showing distribution of neighborhood correlations for each integrated cluster. Only clusters with cells either of positive or negative correlation >80% are shown. Right: Bar plots showing cell-type composition of each cluster in RA (left) and JIA (right) synovium. For each cluster, the most abundant original cell type (from either RA or JIA) is annotated with text in each bar. Center bar plot showing relative proportion of cells derived from JIA versus RA within each cluster (JIA in orange, RA in blue). **e**, Box plots showing the lineage-normalized proportion of cell clusters across individual samples, stratified by condition (JIA vs RA synovium). *P*-values were calculated using Wilcoxon rank-sum tests. Box plots showing the median, interquartile range, and 1.5× interquartile range (IQR) whiskers. **f**, Illustration summarizing key cellular interactions and spatial organization observed in JIA synovium. Pro-inflammatory macrophages interact with CD8^+^ T cells, the resource of *IFNG*, and CD4^+^ T cells, via the CXCL9–CXCR3 chemokine axis. In the perivascular region, *NOTCH3*^+^ sublining fibroblasts engage in Notch signaling with endothelial cells. *MERTK*^+^ macrophages are distributed throughout the sublining area, with local enrichment around endothelial cells. TLS were composed of CD4^+^ T cells, CD8^+^ T cells, B cells, plasma cells, and mregDCs.

Unsupervised clustering in the integrated latent space yielded 42 shared cell clusters encompassing cells from both JIA and RA samples (**Fig. 6c**). To identify disease-relevant cell states, we applied a CNA approach. This approach identified clusters with significantly different neighborhood compositions between the two conditions; for instance, integrated-clusters 10 was expanded in JIA, while integrated-clusters 21 was depleted (**Fig. 6d**).

To mitigate potential technical artifacts arising from differences in tissue dissociation sensitivity between spatial transcriptomics and scRNA-seq(59), we performed lineage-normalized comparisons between JIA and RA by calculating the relative abundance of key subsets within major immune lineages, with focus on cell populations which were dominant in skewed integrated cell clusters toward JIA or RA (**Fig. 6e**). Specifically, *NOTCH3*^+^ sublining fibroblasts (matched with mural cells in the RA-cluster), *CXCL12*^+^ sublining fibroblast cells, and *GZMK*^+^ CD8^+^ T cells were enriched in JIA, whereas clusters enriched in RA included *GZMB*^+^ CD8 T cells. Although it did not show statistical significance, Tph/Tfh cells showed a trend toward enrichment in RA (p=0.089). Collectively, these findings demonstrate the power of integrated spatial-single cell analysis to reveal potential cellular distinctions between JIA and RA pathogenesis.

## DISCUSSION

In this study, to our knowledge, we present the largest subcellular-resolution spatial transcriptomic atlas of JIA synovium. We profiled whole synovial sections with single-molecule transcript detection, capturing a rich landscape of diverse immune and stromal cell populations in situ. By mapping cells in their native tissue context, our atlas reveals how innate and adaptive immune cells, fibroblasts, and endothelial cells organize into discrete pathogenic niches within the JIA joint. Key inflammatory cell states identified (e.g. pro-inflammatory macrophages, Tph/Tfh, *GZMK*^+^/*GZMB*^+^ CD8^+^ T cells, innate-like T cells, B/plasma cells, and distinct fibroblast subsets) spatially co-localize in patterns consistent with functional crosstalk (**Fig. 6f**).

The identification of spatially resolved tissue niches within JIA synovium reveals distinct immune–stromal architectures that likely underpin localized pathogenic processes. Our unsupervised classification of microanatomical niches into four major classes—T+B/plasma cell, Myeloid+Stromal, Myeloid+T cell, and Stromal—reflects the complex cellular crosstalk shaping the inflamed synovial landscape. The Myeloid+T cell and T+B/plasma cell niches exhibited enrichment for IFN-α/γ signaling and adaptive immune pathways, respectively, echoing findings from adult RA studies in which Th1-type cytokines and B cell–T cell interactions dominate lymphoid-rich synovitis(28). The presence of angiogenesis-associated signatures within stromal-rich niches further supports prior work suggesting that fibroblast cells with enrichment for genes associated with the vasculogenesis program sustain chronic inflammation(60, 61). These spatially confined signaling environments may facilitate niche-specific therapeutic resistance or responsiveness, as prior study has shown that pauci-immune synovium with abundant fibroblasts is associated with poor therapeutic response in RA(28, 62). Future studies integrating spatial proteomics and longitudinal biopsy cohorts could clarify how these niches evolve over disease course or in response to therapy, ultimately advancing precision medicine strategies in JIA.

Synovial fibroblasts are broadly classified into lining and sublining fibroblasts: sublining fibroblasts exhibit a secretory phenotype characterized by high expression of inflammatory molecules such as IL-6, CXCL12, and CCL2 which recruit monocytes, whereas lining fibroblasts are the source of the metalloproteinase as the main driver of invasion and cartilage degradation(63, 64). A striking finding in our study is the spatial coupling of sublining fibroblasts with the synovial endothelium via NOTCH3 signaling, a niche interaction that mirrors observations in RA(8). In RA, endothelium-derived NOTCH signals imprint a positional identity on perivascular fibroblasts, promoting an aggressive inflammatory phenotype(8)￼. The shared endothelial– fibroblast niche architecture in JIA and RA underscores a fundamental pathological circuit and raises the hypothesis that targeting the NOTCH3 pathway could attenuate fibroblast pathogenicity in JIA, echoing therapeutic insights previously shown in RA models(8). Interestingly, when integrating the JIA Xenium spatial transcriptomic data with RA scRNA-seq data, many of the *NOTCH3*^+^ fibroblasts in JIA synovium were predicted to align with mural cell identities, suggesting continuous cell states between them. This finding is congruent with the previous study showing that there are no universal specific markers to distinguish fibroblasts and mural cells(65) and that mural cells can arise from perivascular fibroblast-like cells through NOTCH signaling(66). Furthermore, when we applied gsMap, *PRG4*^+^ lining fibroblasts showed the strongest association with JIA GWAS signals. This finding stands in contrast to the inflammatory sublining fibroblasts enriched in patients with high CRP, and suggests that distinct fibroblast phenotypes may be differentially involved in disease susceptibility versus disease activity or progression. Such a model parallels observations in systemic lupus erythematosus (SLE), where gene programs associated with disease-state differ from those linked to severity(67).

Our spatial analysis also uncovered intimate interactions between infiltrating T cells and synovial macrophages that appear to form a positive feedback loop driving pro-inflammatory-polarized inflammation. In JIA synovium, *CXCR3*^+^ T cells were frequently colocalized with *CXCL9^+^CXCL10^+^* pro-inflammatory macrophages. CXCL9/CXCL10–CXCR3 chemokine axis is known to recruit and activate Th1 cells in inflamed joints(68): synovial macrophages (and lining cells) secrete CXCR3 ligands CXCL9/CXCL10 under IFNγ stimulation(69)￼, which align with enrichment of IFN-α/γ signaling in Myeloid+T cell niches described in our analysis. Our data support this model in situ, suggesting that T cells and pro-inflammatory macrophages engage in reciprocal activation – effector T cells maintains macrophages in an pro-inflammatory state, and macrophage-derived CXCL9/CXCL10 attracts more T cells into the tissue. Importantly, our spatial transcriptomic data further revealed that *GZMB*^+^ CD8^+^ T cells exhibited the highest per-cell expression of *IFNG* transcripts in the inflamed JIA synovium. Although *GZMB*^+^ *CD8*^+^ T cells represented a small fraction (<2%) of total T cells, we demonstrated that CD8*^+^* T cells were the largest source of IFNγ. These findings point to a pathogenic circuit in which a small population of highly activated CD8⁺ T cells exerts a disproportionately large influence on the inflammatory environment through sustained IFNγ production.

*MERTK*^+^ macrophages and *TREM2*^+^ macrophages identified in our study correspond to functional analogs of LYVE-1^+^ perivascular macrophages and CX3CR1^+^ lining macrophages previously described in mouse models, respectively(52, 70). In parallel, RA remission has been associated with *MERTK*^+^ macrophage subsets, including *LYVE1*^+^ and *TREM2*^high^ populations(18). The spatial proximity of *MERTK*^+^ macrophages to endothelial cells, combined with their transcriptional signature, supports a role for these cells in vascular maintenance and resolution of inflammation in the pediatric synovium. In RA, *MERTK^+^LYVE1^+^* macrophages predominantly reside in the lining layer during homeostasis and remission, but shift to perivascular regions in the sublining layer during active inflammation(18). The localization of *MERTK^+^*macrophages in our JIA samples mirrors this pattern and may reflect the inflammatory state of the tissue, as all biopsies in our study were obtained from clinically active joints. Moreover, recent work has highlighted a specialized vascular–immune interface at the synovial lining–sublining boundary, where fenestrated *PV1*^+^ endothelial (proxy for venules in our dataset) cells permit entry of circulating immune complexes, initiating inflammatory responses(46). These are sensed by resident macrophages and nociceptor neurons, which act as components of the synovial defence system, and potential secondary effectors of fibroblasts. The enrichment of endothelial–fibroblast interactions and the anatomical proximity of perivascular macrophage subsets in JIA synovium are consistent with this emerging model of immune surveillance.

Using spatial colocalization analysis, we identified organized aggregates of B cells, T cells, and dendritic cells in the synovium, often with structures reminiscent of germinal centers. These TLS-like aggregates in JIA closely mirror those observed in adult autoimmune diseases such as RA and SLE, amplifying local autoimmunity and inflammation(71, 72)￼. Likewise, JIA TLS may serve as sites of local antigen presentation and lymphocyte activation, potentially driving epitope spreading or autoantibody production even in a pediatric setting, although the specific antigens involved remain unknown. In the context of JIA, lymphoid aggregates in the synovium have been reported(57). Intriguingly, these nascent TLS in JIA did not strongly associate with disease duration or severity, though they were more common in antinuclear antibody (ANA)-positive cases. The TLS we observed shared molecular features with TLS in other disorders – for example, JIA TLS areas showed upregulation of lymphoid chemokines *CXCL13* from Tph/Tfh cells and *CCL19* from mregDCs, and *CD40* in B cells(73). These findings encourage further study of TLS as a prognostic marker and as a potential therapeutic target, especially given that some of the agents targeting TLS-related pathways are currently in clinical trials for RA(74).

While JIA and RA are distinct diseases(1), our findings might enable a cross-comparison of pediatric vs. adult synovial pathogenesis. Within stromal cells, the cellular composition of inflamed JIA synovium skewed toward inflammatory elements, including *NOTCH3*^+^ and *CXCL12*^+^ sublining fibroblasts compared to RA. The preferential enrichment of *GZMK^+^* CD8*^+^*T cells in JIA and *GZMB^+^* CD8^+^ T cells in RA suggests that divergent modes of tissue inflammation and immune activation via GZMK and GZMB pathways(75, 76) may contribute to different pathogenic mechanisms in pediatric versus adult arthritis. Furthermore, they may reflect differences between seronegative JIA (RF-negative) and seropositive RA, the latter of which includes the majority of RA patients in our comparison, with over 75% testing positive for anti-cyclic citrullinated peptide antibodies. Nonetheless, these contrasts between adult and pediatric arthritis, although preliminary, provide hypotheses about how age and immune maturation influence the inflammatory microenvironment.

Methodologically, this study introduces a new computational pipeline for quantifying spatial colocalization at both cell-type and single-spot levels. The latter approach yielded quantitative “colocalization scores” for cell pairings and enabled linking with the established framework for inferring ligand–receptor signals. While prior single-cell studies in RA inferred cell interactions from dissociated cells(77–80)￼, our pipeline provides a spatially resolved view of cell communications, adding confidence by requiring physical proximity in tissue. We envision that this analytic framework can be applied to other spatial omics datasets, enabling a deeper understanding of tissue organization and cross-talk in health and disease.

This work has several limitations that warrant discussion. First, the number of patient samples is modest, reflecting the difficulty of obtaining pediatric synovial biopsies; thus, the generalizability of some findings (e.g. TLS presence or specific rare cell populations) will need confirmation in larger JIA cohorts. Relatedly, while we systematically tested associations between all cell populations and multiple clinical covariates—including disease duration, treatment status (e.g., MTX, bDMARDs, steroid injection), and the presence of uveitis—we found no statistically significant associations apart from CRP levels, underscoring the need for validation in larger cohorts to better define the relationship between synovial cell composition and clinical phenotypes. Second, our comparisons between JIA and RA relied on integrating our spatial transcriptomics data with published scRNA-seq data from adult RA, and we acknowledge that such cross-study comparisons are inherently indirect due to differences in tissue processing and assay platforms. However, these differences can also offer complementary insights. For instance, spatial transcriptomics has a higher capture rate of stromal cells compared to dissociative scRNA-seq, which often underrepresents these cell types(59). This allowed us to characterize immune– stromal interactions more comprehensively in JIA synovium.

In conclusion, our high-resolution spatial transcriptomic data of oligo/poly JIA synovium in combination with new computational pipelines charted the inflammatory cell atlas and microanatomy of the disease with unprecedented detail, paving the way for future studies to leverage spatial omics in understanding tissue-level immunology. By combining spatial genomics with multi-omics and experimental validation, we move closer to unraveling the cellular choreography of arthritis and translating these insights into targeted therapies for children and adults suffering from joint inflammation.

## METHODS

### Subject recruitment and clinical data collection

Children who met the International League of Associations for Rheumatology (ILAR) classification criteria for a diagnosis of JIA(3) and were seen at Children’s Hospital Colorado (CHCO) were recruited for the study. Clinical information was obtained through a review of medical records. ANA and RF were considered positive if detected at any titer and at any point during the disease course. CRP levels were measured within three months of the procedure. The design and conduct of this study fully complied with all relevant regulations regarding the use of human study participants and adhered to the ethical principles outlined in the Declaration of Helsinki.

### Collection of sample and processing

Synovial tissue samples were obtained via ultrasound-guided biopsy using a Quick-Core needle. Six to eight tissue fragments were fixed in formalin for subsequent paraffin embedding and histological processing.

FFPE tissue sections were dried in an oven, deparaffinized through a xylene gradient, and de-crosslinked according to the manufacturer’s instructions (10x Genomics protocol CG000578 and CG000580). The Xenium Prime RNA probe set (10x Genomics) was hybridized overnight, ligated, and amplified using rolling circle amplification (10x Genomics protocol CG000760). All incubations are performed using reagents provided by 10x Genomics on a VeritiPro Thermocycler (Applied Biosystems). Autofluorescence was quenched and morphology stains were added, including nuclear, cytoplasmic, and membrane stains for improved cell segmentation (10x Genomics). Slides were then loaded onto the Xenium instrument for whole slide tissue overview scanning and region selection. Regions containing tissue were selected and the onboard cyclic decoding and imaging was initiated over a 72-120-hour period. Data was processed on the instrument after image acquisition, performing probe deconvolution, cell segmentation.

### Histology Assessment

FFPE tissue sections were used for histological analysis. Hematoxylin and eosin (H&E)-stained synovial biopsy sections were examined, and a pathologist assigned Krenn inflammatory infiltrate score.

### Xenium data preprocessing

Raw Xenium datasets were loaded and processed individually using the Seurat v5 R package(81). Data was read using a customized function, capturing gene expression matrices and spatial coordinates, including centroids and segmentations. After loading, each dataset was filtered to retain cells with detected genes between 200 and 5000 and total transcripts less than 25,000. SCTransform was applied individually to each sample for normalization and variance stabilization. Subsequently, the nine samples were merged into a single Seurat object, followed by another round of SCTransform(82) to ensure integrated normalization. We then performed principal component analysis (PCA) with 20 principal components (PCs) on the merged dataset with 2000 variable features and performed Harmony(83) integration (lambda=1, theta=2, and sigma=0.1) to correct for batch effects by different slides and samples. Uniform Manifold Approximation and Projection (UMAP) were applied post-integration. Cell cycle phase was inferred by cell types using CellCycleScoring function in the Seurat R package.

### Cell type identification

We constructed a shared nearest neighbor (SNN) graph based on the first 20 harmonized PCs and identified initial broad cell types based on clustering results with the Louvain algorithm at resolution 0.20. Broad clusters were categorized into (1) stromal tissue cells (fibroblasts and endothelial cells), (2) myeloid cells, (3) T cells/ILCs (including NK cells), and (4) B/plasma cells based on the expression of canonical markers. Cells identified as belonging to these broad cell types were then extracted into separate datasets for further analysis. Each cell type underwent a second round of SCTransform normalization, PCA, Harmony integration, and UMAP dimensionality reduction. Louvain clustering was again performed separately on these subsets, with resolutions tailored specifically for each cell type (T/ILCs: 1.00, B/plasma: 0.40, myeloid: 0.20, stromal tissue: 0.60) to achieve refined subpopulation identification. The effect of batch correction was assessed using Local Inverse Simpson’s Index (LISI)(83). Cell clusters were annotated based on the expression of canonical marker genes. To identify top10 marker genes characteristic of each cluster, we applied the wilcoxauc function from the presto R package(84) to normalized expression data. Differential expression was first assessed between each cluster and all remaining cells using the Wilcoxon rank-sum test. In addition, to account for potential confounding from broad lineage differences, we repeated the differential expression analysis within each cell type, comparing each subcluster to other subclusters. Following differential expression analysis, we selected the top 10 genes per cluster ranked by log fold change and reported in **Supplementary Table 2**.

### Identification of cell populations that are associated with specific clinical variables

To identify cell states with disease-specific spatial organization, we implemented a CNA(85). Briefly, CNA quantifies the relationship between each cell based on transcriptome similarity, allowing detection of transcriptome neighborhoods covary with sample-level conditions such as disease status (e.g., CRP level). We first defined a SNN graph using harmonized PCs. This matrix was then used to calculate a neighborhood correlation score.

### Integration of spatial transcriptomics and GWAS data using gsMap

To identify genetically relevant spatially localized cell types within the synovium, we applied gsMap, a computational framework that integrates spatial transcriptomic profiles with GWAS summary statistics(33). In the gsMap framework, each cell was embedded into a low-dimensional representation that captured both molecular and spatial context. Gene specificity scores were computed for each cell based on the learned latent space, representing how specifically each gene is expressed in a given cell’s spatial context. Then, these gene specificity scores were integrated with GWAS signals for JIA (GWAS catalog: GCST90010715)(34). Using precomputed LD scores and SNP weights from the 1000 Genomes Project European reference panel (Phase 3), gsMap estimated the heritability enrichment for each spatial cell by modeling the association between GWAS summary statistics and spatially defined LD scores.

### Identifying spatial niches

To identify spatial niches, we took two complementary approaches. First, we utilized a method inspired by Goltsev et al.(86) and He et al.(87), implemented in Seurat’s BuildNicheAssay function. This method defines the ‘local neighborhood’ for each cell by considering its ‘k.neighbor’ spatially closest neighbors and counts occurrences of each cell type present within this neighborhood. Cells sharing similar neighborhood compositions are grouped into spatial niches using k-means clustering. We made slight modification of this function to identify corresponding niches across different slides and used k.neighbor=30. To identify biologically interpretable niche clusters within identified niches, we then constructed a cell-type composition matrix across all identified niches. This matrix was derived by calculating the proportion of each annotated cell type in each spatial niche, excluding “mixed”, “proliferating”, and “muscle cell” clusters to reduce noise. PCA was performed on the normalized composition matrix to reduce dimensionality and capture the major sources of variation in niche composition. We determined the optimal number of PCs to retain based on the cumulative proportion of variance explained, selecting the smallest number of PCs that accounted for at least 30% of the total variance, resulting in the top 3 PCs in total for the downstream process. To classify niches into broader biologically meaningful categories, we applied k-means clustering to the PCA-reduced data. The optimal number of clusters was selected based on the average silhouette width, which evaluates the compactness and separation of clusters. The k=4 that maximized the average silhouette score was chosen as the optimal value. Based on the dominant cell types in each cluster and biological interpretability, we manually annotated each niche cluster as one of the following: Stromal niche, Myeloid + Stromal cell niche, Myeloid+T cell niche, or T + B/plasma cell niche. To assess pathway activity within spatially defined niches, we applied the escape R package (v2.2.3)(88) to perform single-cell gene set enrichment analysis(89). We used a curated collection of gene sets from the Molecular Signatures Database (MSigDB)(90), including Hallmark gene sets(91) and KEGG pathway gene sets(31). Gene sets were filtered to retain only those genes expressed in the dataset. Enrichment scores were computed on the normalized expression matrix using escape.matrix using “ssGSEA” method(89) with 5000 random background groups and a minimum gene set size of 5. Scores were subsequently normalized using the performNormalization function to account for differences in gene detection rate, ensuring comparability across cells.

Secondly, we developed a custom method that performs spatial neighborhood enrichment analyses. For each slide, spatial coordinates (‘centroids’) of cells were obtained, and the nearest neighbors for each cell were identified using the FindNeighbors function in Seurat with a parameter ‘neighbors.k = 30’. We then constructed an adjacency matrix representing spatial relationships among cells by combining nearest neighbor information across slides using block diagonal matrices. The observed spatial enrichment of cell type interactions was quantified using a custom function, which calculates occurrences of each pairwise cell type interaction within spatial neighborhoods. To assess the statistical significance of these observed interactions, 500 permutation tests were performed by randomly shuffling cluster labels across cells. Z-scores were calculated to measure how significantly the observed neighborhood compositions deviated from randomized expectations.

### Module score calculation for pro-inflammatory macrophage, LYVE-1^+^ perivasuclar macrophage, and CX3CR1^+^ lining macrophage signature

To quantify the pro-inflammatory polarization state in macrophages, we computed a module score based on a curated set of pro-inflammatory marker genes. The gene set was derived from published studies that define transcriptional programs associated with pro-inflammatory signature(49, 50), including *ZBP1*, *RSAD2*, *TAP2*, *VCAM1*, *BATF*, *CD86*, *SERPINE1*, *PVR*, *CXCL16*, *CCND2*, *PDPN*, *CCRL2*, *FLNB*, *MAP3K5*, *IL17RA*, *RBPMS*, *INHBA*, *TRAF1*, *NFKB2*, *VASP*, *PTGES*, *IFNAR2*, *IL1RN*, *CD274*, *NOTCH1*, *IFI35*, *IFIT2*, *ICAM1*, *IL12B*, *IL12A*, *CD14*, *ITGA5*, *EIF2AK2*, *PILRA*, *CFLAR*, *TNIP1*, *TNFRSF1B*, *TLR2*, *MMP14*, *OAS3*, *ADORA2A*, *JDP2*, *IRF7*, *MITD1*, *CD40*, *CXCL9*, *GRAMD1A*, *EBI3*, *SOCS3*, *PSTPIP2*, *ACP5*, *CD38*, *IL15RA*, *SERPINB2*, *STAT1*, *ARG2*, *SYK*, *GCH1*, *STAT2*, *HCK*, *MET*, *HDC*, *SNX10*, *ITGAL*, *PTGS2*, *IFIH1*, *TRIM25*, and *JAK2*. For each cell, we calculated the average expression level of the pro-inflammatory marker genes using the row-wise mean across the normalized expression matrix.

In the same way, we computed a module score for the LYVE-1^+^ signature using genes upregulated in FOLR^high^LYVE-1^+^ macrophages, listed in Supplementary Data 1 of the original publication(18). Genes not detected in our dataset were excluded. The final FOLR^high^LYVE-1^+^ macrophage gene set used for scoring included the following genes detected in our dataset: *F13A1, SLC40A1, STAB1, PLTP, FOLR2, LGMN, HMOX1, COLEC12, GAS6, MRC1, DAB2, MAF, THBD, EMP1, LILRB5, CTSC, PMP22, PEPD, SLCO2B1, GPR34, CSF1R, TSPAN4, CD14, NINJ1, CD4, MEF2C, RBPJ, MAFB, IFI16, CLTC, ABL2, CD59, CTSL, EIF4E, FCGR2B, ATF3, CD99, UCP2, LAMP1,* and *FCGR2A*.

To investigate the presence of barrier-forming macrophage phenotypes in our dataset, we curated a gene signature associated with CX3CR1^+^ lining macrophages, as reported by Culemann et al.(52). The signature was derived from the set of marker genes enriched in CX3CR1^+^ macrophages (upregulated genes in lining vs. interstitial macrophage in Fig. 3B and marker genes of cluster 3 in Fig. 3C-D of the original paper), and corresponding mouse gene symbols were converted to human orthologs using Ensembl annotation and manual curation. After intersecting with our measured genes, a module score was calculated using the following genes: *FAT3, CX3CR1, ADCYAP1R1, CDH23, ALDOA, FABP7, HTR2B, SIRPB1, TSPAN18, NPNT, OLFML3, and VSIG4*.

### Simulation analysis for method evaluation

To evaluate the performance of our spatial neighborhood enrichment analyses, we employed simulation data generated using custom R scripts. Two spatial configurations, termed “concentric circle” and “layer”, were simulated to assess the sensitivity and robustness of our method in detecting predefined spatial neighborhood patterns. For each simulation scenario, we generated datasets comprising 1,500 cells randomly distributed across a defined spatial domain (800 × 800 μm). Cells were assigned into 10 distinct cell types. To create spatially enriched interactions, we specifically arranged two cell types to exhibit spatial proximity while randomly distributing the remaining cell types within the spatial field.

In the concentric circle scenario, cells belonging to the anchor cell type (reference) were primarily concentrated within a central circular area, whereas neighboring cells were positioned to form an outer ring surrounding this central region at a fixed distance. Additional random noise was introduced to mimic realistic spatial variation. Cells from other cell types were uniformly randomly distributed across the entire spatial domain.

In the layer scenario, spatial enrichment was structured along layer patterns. Reference cells were positioned along a straight horizontal line, with neighboring cells located in parallel at a fixed vertical offset. As with the circle scenario, random spatial noise was added to replicate biological heterogeneity.

Using these synthetic spatial distributions, we performed neighborhood enrichment analyses to validate the sensitivity of our method to detect known spatial relationships. We then assessed spatial enrichment of cell type interactions by computing z-scores from 500 permutations by randomly shuffling cluster labels across cells. Significant cell-cell enrichments were determined based on Benjamini-Hochberg FDR-corrected p-values.

### Colocalization score

To investigate spatial proximity at the single-cell/spot level, we computed a colocalization score that measures the local co-occurrence of two specified cell clusters. This analysis complements the spatial neighborhood approach by providing insights at the resolution of individual cells rather than at the broader cell-type level. For each cell, we first identified neighboring cells within a defined radius using spatial coordinates derived from imaging data. Specifically, we utilized the nn2 function in the RANN R package(92) to find neighbors around a cell of the anchor cell type within a radius of 10 µm. We then assessed whether both anchor and target clusters were present among these neighboring cells. A binary score was assigned to each cell, indicating the simultaneous presence (score of 1) or absence (score of 0) of both clusters within its immediate spatial neighborhood. To account for broader spatial context and to smooth local colocalization signals across the tissue, we implemented a random walk, inspired by covarying neighborhood analysis(85), based on the adjacency matrix derived from the spatial neighbor network. The adjacency matrix was constructed using spatial coordinates from multiple samples, integrated using a block-diagonal matrix. In the random walk step, the local colocalization scores were propagated iteratively through neighboring cells, weighted by their spatial connectivity, until convergence was determined by monitoring changes in the kurtosis of the score distribution. This iterative diffusion process was limited to a maximum of 15 steps to ensure computational efficiency while achieving stable estimates of spatial co-occurrence patterns. The final result provided a quantitative measure for each spatial spot, reflecting its degree of spatial co-occurrence with the specified clusters in its immediate neighborhood, allowing precise identification of spatially defined microenvironments and potential cell-cell interactions.

### Public spatial transcriptomic dataset acquisition and preprocessing

Public spatial transcriptomic datasets from 10X Genomics Xenium platform were utilized for benchmarking purposes. The human breast dataset and mouse brain dataset were downloaded from the 10X Genomics website and preprocessed using the Seurat framework.

For each dataset, spatial gene expression matrices and spatial coordinate metadata were imported. Cells with zero total gene counts were excluded from downstream analysis. Gene expression values were normalized using SCTransform and PCA was performed. SNN graphs were constructed, and clustering was performed using the Louvain algorithm with resolution at 0.6. For the human breast dataset, cell types were annotated based on cluster-specific marker gene expression derived using FindAllMarkers function, followed by manual inspection and comparison with known markers for each tissue. The mouse brain dataset was annotated, following instruction in the Seurat tutorial, using the RCTD (Robust Cell Type Decomposition) pipeline(93) with a reference atlas of Allen Brain subclass annotations downloaded from https://www.dropbox.com/s/cuowvm4vrf65pvq/allen_cortex.rds?dl=1. The spatial object was processed with create.RCTD() and run.RCTD() in “doublet” mode, and predicted cell types were assigned to each spot based on highest posterior probability.

We applied a spatial neighborhood analysis approach to evaluate non-random co-occurrence of cell types. We computed a Z-score-based enrichment matrix from observed cell-type neighbor frequencies compared to a null distribution of 500 spatial permutations. Neighborhoods were defined by k-nearest neighbors (k = 30). To assess spatial colocalization score between specific cell types at the single-spot level, we used a search radius of 30 μm and up to 15 neighborhood expansion steps.

### Ligand–receptor analysis

To systematically identify ligand–receptor interactions at single-spot resolution, we performed ligand–receptor analyses by samples using COMMOT(54), a Python-based toolkit designed for spatial communication inference. COMMOT integrates spatial coordinates and transcriptomic data to infer ligand–receptor signaling interactions between spots. An integrated ligand–receptor database was constructed by combining human ligand–receptor pairs from CellChat(77) and CellPhoneDB v4.0 databases(77, 78). Spatial ligand–receptor communication was computed using the spatial_communication function with the merged database. We set the spatial distance threshold to 200 μm and enabled analysis of heteromeric ligand–receptor complexes. COMMOT analysis generated spatial communication scores indicating the strength of ligand–receptor interactions from sender and receiver perspectives for each spatial spot.

### Flow cytometry and intracellular cytokine staining

Synovial fluid mononuclear cells were isolated from patients with oligo/poly JIA using Ficoll-Paque density gradient centrifugation. Cells were resuspended in complete RPMI medium and stimulated for 6 hours at 37°C with 50 ng/mL PMA and 1 µg/mL ionomycin (Biolegend), 10 ng/mL IL-12 (200-12, PEPRO-TECH), 50 ng/mL IL-15 (200-15, PEPRO-TECH) and 50 ng/mL IL-18 (B001-5, MBL) with last 4 hours in the presence of 10 µg/mL brefeldin A (BioLegend). After stimulation, cells were washed, stained with Zombie NIR Fixable viability dye (Biolegend), and surface-stained with fluorochrome-conjugated antibodies against CD3 (clone OKT3, Biolegend), CD4 (clone SK3, Biolegend), CD8 (clone SK1, Biolegend), CD45 (clone 2D1, Biolegend), CD56 (clone 5.1.H11, Biolegend), and TCRγδ (clone 3C10, Biolegend). Cells were then fixed and permeabilized using the Cytofix/Cytoperm kit (BD Biosciences) and stained intracellularly with anti–IFN-γ antibody. Flow cytometry was performed on a Cytek Aurora and data were analyzed using FlowJo v10. All antibodies were diluted 1 to 100.

### Identification of tertiary lymphoid structures

To systematically identify TLS, we developed a custom pipeline leveraging the colocalization score described above. First, we calculated the colocalization scores specifically between broad B-cell and T-cell populations. Next, we used spatial coordinates (centroids) obtained from each slide to construct spatial adjacency graphs for individual cells with positive colocalization scores for B-cell and T-cell populations, enabling a more efficient search for relevant interactions compared to scanning the entire spatial transcriptomic dataset. For this step, nearest neighbor cells were identified using a radius-based nearest neighbor search, the nn2 function in the RANN R package, with parameters set to a radius of 50 μm and the maximum number of nearest neighbors set to 200. Subsequently, an undirected spatial adjacency graph was constructed where each node represents a cell, and edges connect spatially proximate cells identified in the radius-based search. Using the igraph package, connected components within this graph were identified, each representing clusters of spatially associated cells. Clusters meeting predefined thresholds—specifically, those containing at least 20 B cells and T cells—were annotated as candidate TLS. The resulting candidate TLS identified through this computational pipeline were subsequently validated by a single pathologist who was blinded to computational analysis results.

To investigate the spatial organization of immune cells relative to B cells within TLS, we calculated the Euclidean distance to the nearest B cell for each cell within a TLS region. These distances were binned in 5 μm intervals from 0 to 300 μm. Within each bin, we computed the proportion of cells for each annotated immune cluster relative to the total number of cells in that cluster.

### Immunohistochemistry staining

An antigen retrieval was performed using a Decloaking Chamber (Biocare Medical) at 95 degrees for 1 hour in Tris/EDTA antigen retrieval buffer (Agilent). They were then stained consecutively with primary antibodies for MERTK (clone ab52968, Abcam), TREM2 (clone TREM2/7210, Invitrogen), CD68 (clone KP1, DAKO), CD3E (clone LN10, Leica), CD8 (clone C8/144B, DAKO), and IL1B (clone 3A6, Cell Signaling Technologies) using cyclic multispectral fluorescence immunohistochemistry. Briefly, the slides were blocked, incubated with primary antibodies, followed by horseradish peroxidase (HRP)-conjugated secondary antibody polymer, and HRP-reactive OPAL fluorescent reagents using a Bond RX autostainer (Leica). The slides were stripped between each stain with heat treatment in an antigen retrieval buffer. Spectral references and unstained control images were measured and inForm software v3.0 was used to create a multispectral library reference. Whole slide scans were collected using the 20x objective with a 0.5 μm resolution and were spectrally unmixed with PhenoImager HT v2.0.0 software.

### Comparison with RA CITE-seq data

Cellular Indexing of Transcriptomes and Epitopes by sequencing (CITE-seq) data from RA synovial tissue were retrieved from previously published dataset(28). The dataset was integrated with the Xenium-derived JIA data using the StabMap method(94). Integration was based on variable mRNA features in the JIA Xenium dataset and present in the RA CITE-seq dataset.

UMAP embedding was performed on the integrated PCA. Cell-type labels were transferred from the RA CITE-seq data to the JIA Xenium data through k-nearest neighbors (k-NN) classification, training the model on the RA dataset with known subtype labels. This integration process was carried out independently by each broad cell type. As for cell clusters defined by integrated PCA, only clusters with sufficient cell numbers (n ≥ 300 cells) were considered for downstream interpretation.

## Statistics

The statistical tests performed are indicated in the figure legends or Methods. For the analysis of our data, the Wilcoxon rank-sum test was used for comparing continuous variables between two groups. Spearman’s rank correlation was utilized to examine the relationship between two continuous variables. We corrected for multiple comparisons using the Benjamini-Hochberg procedure to control the false discovery rate. Adjusted *p* values less than 0.05 were considered significant.

## Acknowledgments

We thank Ryan Oakes and Ana Aristiguieta for their invaluable assistance in obtaining patient consent, which was essential for the successful completion of this study. We acknowledge the Human Immune Monitoring Shared Resource (RRID:SCR_021985) and the University of Colorado Cancer Center (P30CA046934) for performing multi-immunofluorescence histochemistry using the Akoya Biosciences Polaris and spatial transcriptomics using the 10x Genomics Xenium platforms. We acknowledge the Accelerating Medicines Partnership (AMP): Rheumatoid Arthritis and Systemic Lupus Erythematosus (AMP RA/SLE) Network for providing the data used in this study, specifically the RA CITE-seq dataset (SynID: syn52297840, Release 1.5/V7). Access to this controlled dataset was obtained under a data use agreement complying with ethical guidelines to protect participant confidentiality. Investigators accessing these data complied with all data access terms and conditions outlined by AMP RA/SLE and Synapse, and the data were not redistributed in any form.

## Author contributions

K.Y., R.F., N.R., H.L., K.H. and C.L. recruited patients, obtained samples, and curated clinical data. R.F., C.L. and K.H. contributed to the procurement and processing of samples and study design. A.H.J. provided disease immunology inputs. M.R.C. performed pathological evaluation of histological images. J.I. led the computational and statistical analyses. J.I. and K.Y. interpreted the results, wrote the manuscript. All authors participated in editing the final manuscript.

## Study approval

This study was approved by the Institutional Review Board at University of Colorado School of Medicine (protocol number 23-2268).

## Data availability

All raw and processed data will be available upon acceptance. Spatial transcriptome data for benchmarking spatial neighborhood analysis and colocalization score was downloaded from https://www.10xgenomics.com/jp/support/software/xenium-onboard-analysis/latest/resources/xenium-example-data. Newly developed R package for spatial neighborhood enrichment analysis and colocalization scoring system and vignette using simulation data and public 10X spatial transcriptome data will be available on Github upon acceptance. Other analytic codes to generate figures will be available on Github upon acceptance.

## List of Supplementary Materials

Supplementary Table 1: Characteristics of enrolled patients.

Supplementary Table 2: Top 10 differentially expressed genes for each identified cell cluster. Marker genes were identified using the Wilcoxon rank-sum test via the wilcoxauc function from the *presto* R package. Analyses were performed both across all cells and within each broad lineage (T cells/ILCs, B/plasma cells, myeloid cells, and stromal tissue cells) to identify cluster-specific markers independent of major cell type.

Supplemental Figures are available in the separated file.

