## Supplemental Figure for "Subcellular spatial transcriptomics reveals immune–stromal crosstalk within the synovium of patients with juvenile idiopathic arthritis"

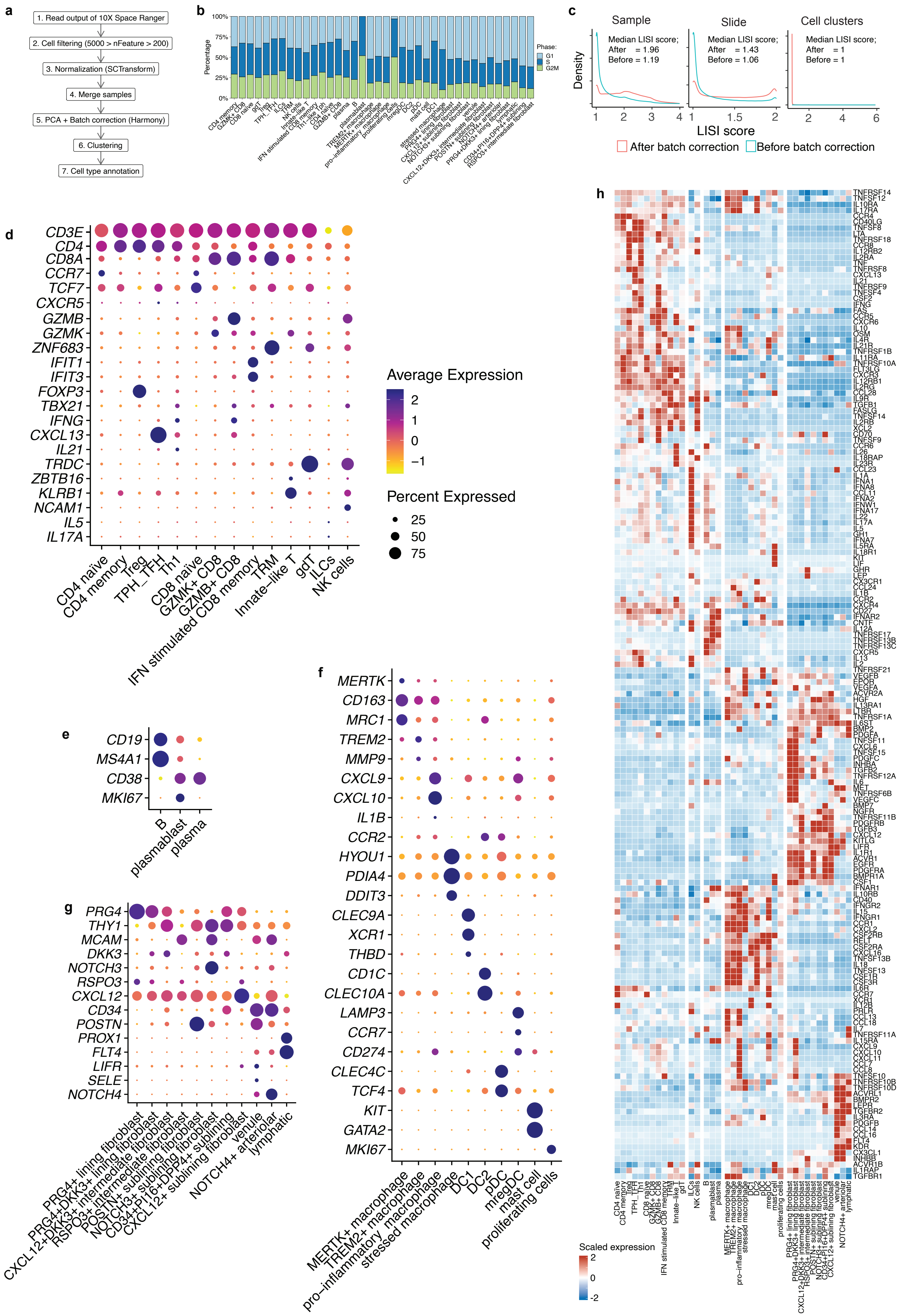
a
1. Read output of 10X Space Ranger
2. Cell filtering (5000 > nFeature > 200)
3. Normalization (SCTransform)
4. Merge samples
5. PCA + Batch correction (Harmony)
6. Clustering
7. Cell type annotation
b
Percentage
100%
75%
50%
25%
0%
Phase:
G1
S
G2M
c
Sample
Median LISI score;
After = 1.96
Before = 1.19
Slide
Median LISI score;
After = 1.43
Before = 1.06
Cell clusters
Median LISI score;
After = 1
Before = 1
Density
LISI score
After batch correction
Before batch correction
d
CD3E
CD4
CD8A
CCR7
TCF7
CXCR5
GZMB
GZMK
ZNF683
IFIT1
IFIT3
FOXP3
TBX21
IFNG
CXCL13
IL21
TRDC
ZBTB16
KLRB1
NCAM1
IL5
IL17A
CD4 naïve
CD4 memory
Treg
TPH_TFH
Th1
CD8 naïve
GZMK+ CD8
GZMB+ CD8
IFN stimulated CD8 memory
TRM
Innate-like T
gdT
ILCs
NK cells
Average Expression
2
1
0
−1
Percent Expressed
25
50
75
e
CD19
MS4A1
CD38
MKI67
B
plasmablast
plasma
f
MERTK
CD163
MRC1
TREM2
MMP9
CXCL9
CXCL10
IL1B
CCR2
HYOU1
PDIA4
DDIT3
CLEC9A
XCR1
THBD
CD1C
CLEC10A
LAMP3
CCR7
CD274
CLEC4C
TCF4
KIT
GATA2
MKI67
MERTK+ macrophage
TREM2+ macrophage
pro−inflammatory macrophage
stressed macrophage
DC1
DC2
pDC
mregDC
mast cell
proliferating cells
g
PRG4
THY1
MCAM
DKK3
NOTCH3
RSPO3
CXCL12
CD34
POSTN
PROX1
FLT4
LIFR
SELE
NOTCH4
PRG4+ lining fibroblast
PRG4+DKK3+ lining fibroblast
CXCL12+DKK3+ intermediate fibroblast
RSPO3+ intermediate fibroblast
POSTN+ sublining fibroblast
NOTCH3+ sublining fibroblast
CD34+P16+DPP4+ sublining
CXCL12+ sublining fibroblast
NOTCH4+ arteriolar
venule
lymphatic
h
Scaled expression
2
0
-2

**Supplemental Figure 1: Preprocessing steps for spatial transcriptome data. a**, Workflow of preprocessing steps. **b**, Inferred cell cycle phase. **c**, Local Inverse Simpson's Index (LISI) scores to measure mixture levels on samples, slides, and cell clusters. After batch effect correction, the mixture level of samples and slides are significantly reduced compared to before correction (Wilcoxon test $p < 0.01$). **d-g,** Dot plot showing marker gene expressions in identified cell clusters, for T/ILC cluster (**d**), B/plasma cluster (**e**), Myeloid cluster (**f**), and stromal cluster (**g**). **h**, Heatmap showing Z-scored pseudo-bulk expression profiles of cytokine and cytokine receptor genes across 38 synovial cell states. Genes were selected from the MSigDB C2 curated collection (M9809). Only genes detected in more than 3% of cells within any cluster were retained (n = 168). Expression values were aggregated at the cluster level and row-scaled to highlight relative expression patterns across cell populations.

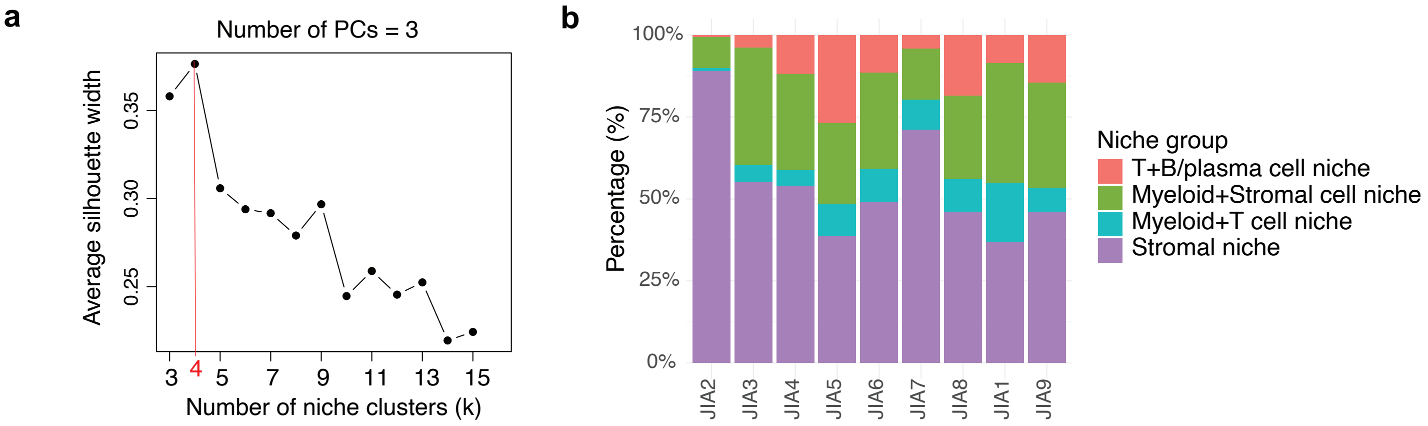

a
Number of PCs = 3
Average silhouette width
0.25
0.30
0.35
3
4
5
7
9
11
13
15
Number of niche clusters (k)
b
Percentage (%)
100%
75%
50%
25%
0%
JIA2
JIA3
JIA4
JIA5
JIA6
JIA7
JIA8
JIA1
JIA9
Niche group
T+B/plasma cell niche
Myeloid+Stromal cell niche
Myeloid+T cell niche
Stromal niche

**Supplemental Figure 2: Identification and distribution of niche clusters across JIA synovial tissues. a,** Silhouette analysis for determining the optimal number of niche clusters. Based on PCA of the niche × cell type composition matrix, the silhouette width was calculated across a range of cluster numbers (k = 3 to 15). The optimal number of clusters was determined to be k = 4 (red line), based on the maximum average silhouette width, using the first 3 principal components (explaining ≥30% variance). **b,** Proportion of each annotated niche group across individual JIA samples. Each bar represents the percentage composition of niche groups within a single patient sample.

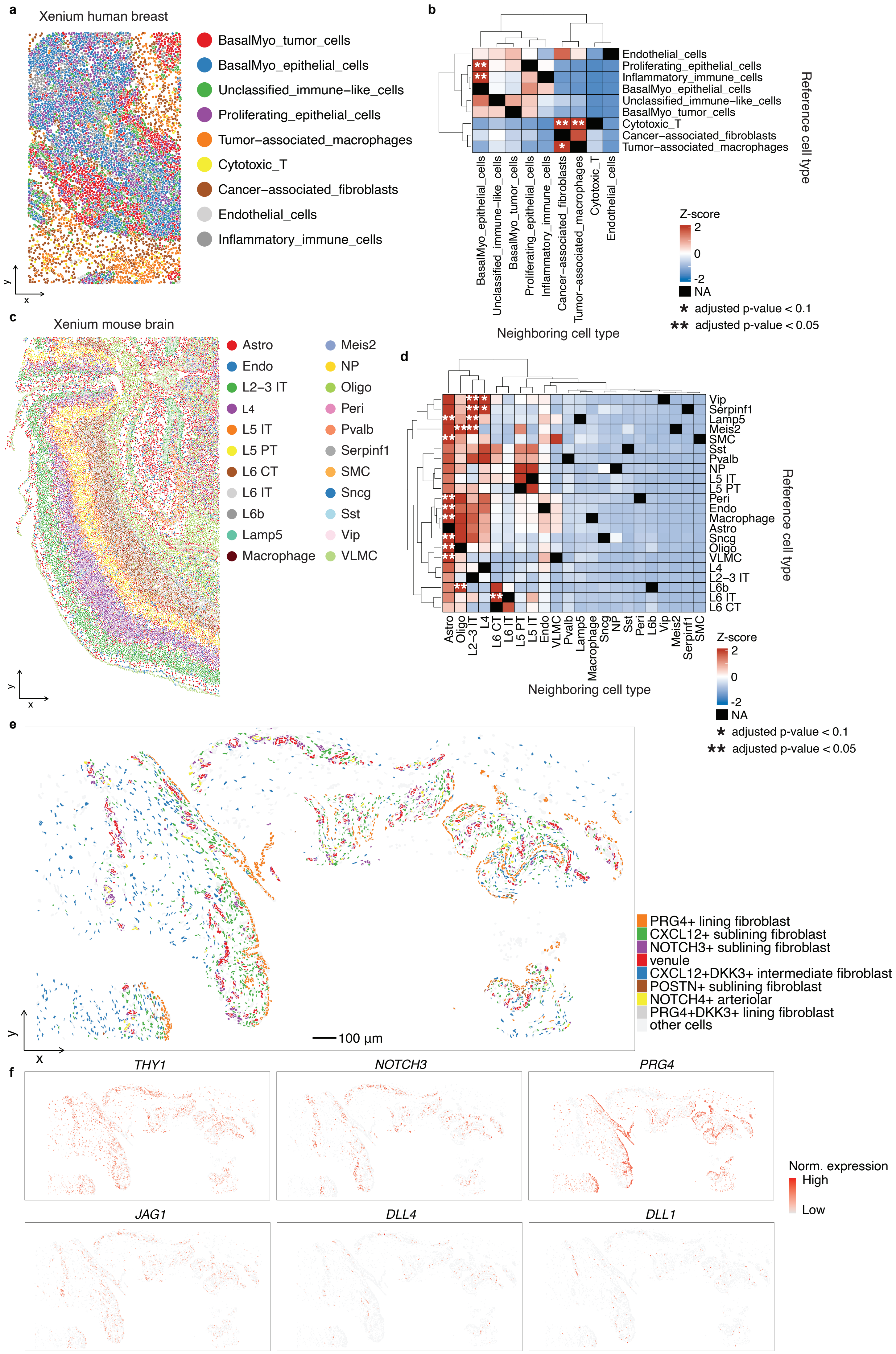
a
Xenium human breast
BasalMyo_tumor_cells
BasalMyo_epithelial_cells
Unclassified_immune–like_cells
Proliferating_epithelial_cells
Tumor–associated_macrophages
Cytotoxic_T
Cancer–associated_fibroblasts
Endothelial_cells
Inflammatory_immune_cells
b
Reference cell type
Neighboring cell type
Z-score
NA
* adjusted p-value < 0.1
** adjusted p-value < 0.05
c
Xenium mouse brain
Astro
Endo
L2–3 IT
L4
L5 IT
L5 PT
L6 CT
L6 IT
L6b
Lamp5
Macrophage
Meis2
NP
Oligo
Peri
Pvalb
Serpinf1
SMC
Sncg
Sst
Vip
VLMC
d
Reference cell type
Neighboring cell type
Z-score
NA
* adjusted p-value < 0.1
** adjusted p-value < 0.05
e
PRG4+ lining fibroblast
CXCL12+ sublining fibroblast
NOTCH3+ sublining fibroblast
venule
CXCL12+DKK3+ intermediate fibroblast
POSTN+ sublining fibroblast
NOTCH4+ arteriolar
PRG4+DKK3+ lining fibroblast
other cells
100 µm
f
THY1
NOTCH3
PRG4
JAG1
DLL4
DLL1
Norm. expression
High
Low

**Supplemental Figure 3: Benchmarking the pipeline of spatial neighborhood enrichment analysis using public spatial transcriptome datasets. a**, Spatial distribution of annotated cell types within the human breast cancer tissue dataset derived from the Xenium spatial transcriptome technology. Each color corresponds to a distinct cell type. **b**, Heatmap of spatial neighborhood enrichment in the human breast cancer tissue. The color gradient represents z-scores indicating significant spatial interactions between pairs of cell types, with red denoting enriched (positively associated) spatial proximity and blue representing depleted (negatively associated) interactions. Statistical significance is annotated as * (adjusted p-value < 0.1 by Benjamini-Hochberg method) and ** (adjusted p-value < 0.05). **c**, Spatial distribution of annotated cell types within the mouse brain tissue dataset derived from the Xenium spatial transcriptome technology. Each color corresponds to a distinct cell type. **d**, Heatmap of spatial neighborhood enrichment in the mouse brain tissue. **e**, Representative spatial coordinates of a identified niche of sub-lining fibroblasts and endothelial cells, matching pathological images. Cells are colored according to their cluster assignments. **f**. Spatial expression patterns of fibroblast-related and Notch signaling-related genes within the identified niche location.

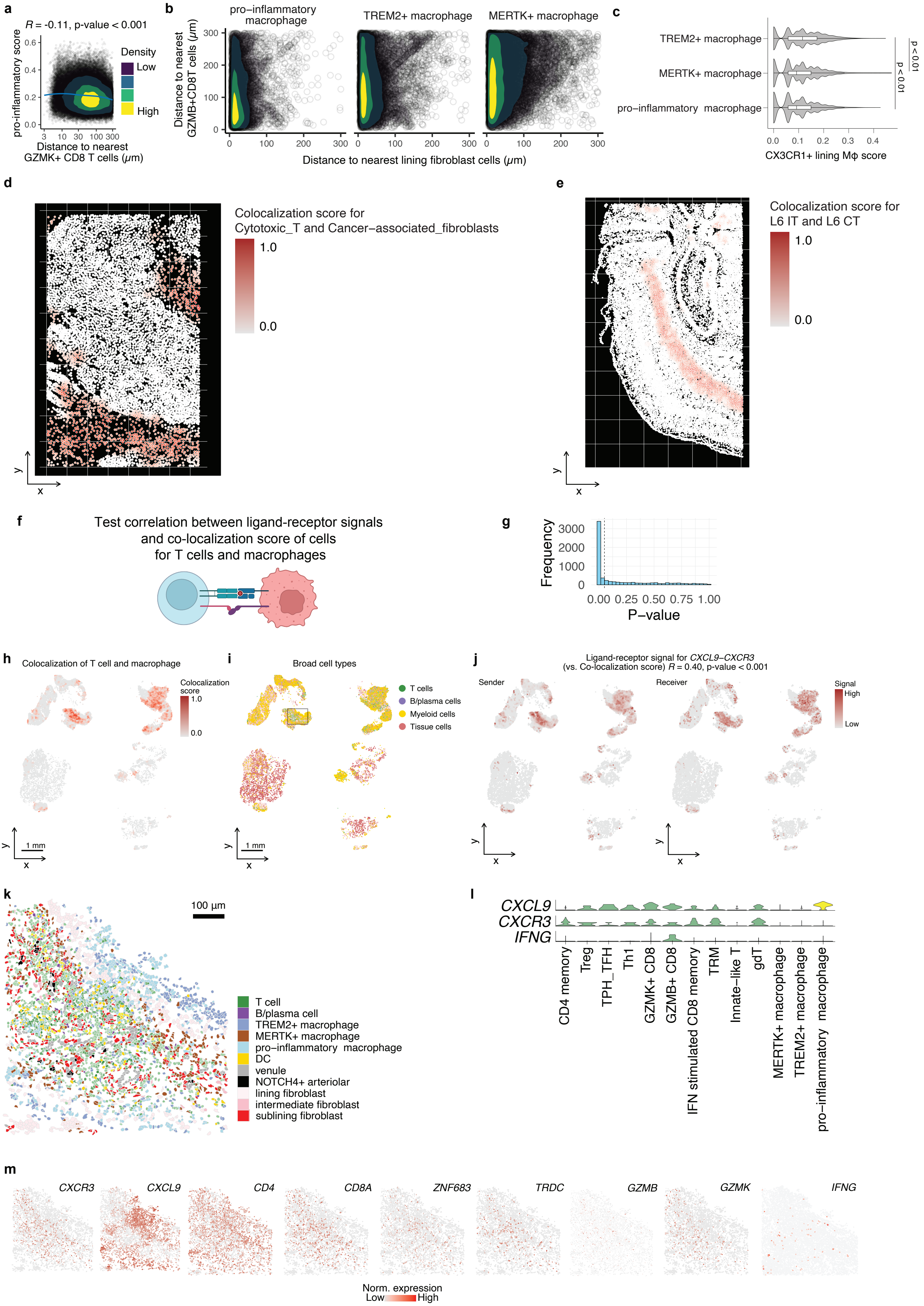
a
R = -0.11, p-value < 0.001
pro-inflammatory score
Density
Low
High
Distance to nearest
GZMK+ CD8 T cells (μm)
b
pro−inflammatory
macrophage
TREM2+ macrophage
MERTK+ macrophage
Distance to nearest
GZMB+CD8T cells (μm)
Distance to nearest lining fibroblast cells (μm)
c
TREM2+ macrophage
MERTK+ macrophage
pro−inflammatory macrophage
p < 0.01
p < 0.01
CX3CR1+ lining Mϕ score
d
Colocalization score for
Cytotoxic_T and Cancer−associated_fibroblasts
1.0
0.0
e
Colocalization score for
L6 IT and L6 CT
1.0
0.0
f
Test correlation between ligand-receptor signals
and co-localization score of cells
for T cells and macrophages
g
Frequency
P−value
h
Colocalization of T cell and macrophage
Colocalization score
1 mm
i
Broad cell types
T cells
B/plasma cells
Myeloid cells
Tissue cells
1 mm
j
Ligand-receptor signal for CXCL9–CXCR3
(vs. Co-localization score) R = 0.40, p-value < 0.001
Sender
Receiver
Signal
High
Low
k
100 µm
T cell
B/plasma cell
TREM2+ macrophage
MERTK+ macrophage
pro−inflammatory macrophage
DC
venule
NOTCH4+ arteriolar
lining fibroblast
intermediate fibroblast
sublining fibroblast
l
CXCL9
CXCR3
IFNG
CD4 memory
Treg
TPH_TFH
Th1
GZMK+ CD8
GZMB+ CD8
IFN stimulated CD8 memory
TRM
Innate-like T
gdT
MERTK+ macrophage
TREM2+ macrophage
pro-inflammatory macrophage
m
CXCR3
CXCL9
CD4
CD8A
ZNF683
TRDC
GZMB
GZMK
IFNG
Norm. expression
Low
High

**Supplemental Figure 4: Benchmarking the pipeline of colocalization scoring analysis using public spatial transcriptome datasets. A,** Correlation between pro-inflammatory module scores of individual macrophage cells (each point) (y-axis) and distance to the *GZMK*+ CD8 T cells (x-axis). Color-filled contours represent two-dimensional kernel density estimates. Spearman's correlation coefficient and p-values are shown. **b,** Density plots depict the distances of individual macrophage cells (each point) to the nearest lining fibroblast cells (x-axis) and *GZMB*+ CD8 T cells (y-axis) by macrophage subtypes. Color-filled contours represent two-dimensional kernel density estimates. **c**, Violin plots showing the distribution of CX3CR1+ lining macrophage module scores across macrophage subtypes. **d-e**, Colocalization scores distribution between Cytotoxic T cells and Cancer-associated fibroblasts in the human breast cancer (**d**) and L6 IT and L6CT in the mouse brain (**e**) spatial transcriptome datasets which were introduced in Supplemental Figure 3. **f**, Shema of the analytical approach used to correlate ligand–receptor interactions with spatial colocalization scores for T cells and macrophages. **g**, Distribution of p-values from Spearman's correlation tests between ligand–receptor signaling strength and colocalization scores between macrophages and T cells. **h**, Spatial heatmap of colocalization scores between T cells and macrophages. **i**, Spatial map of broad cell type annotation including T cells, myeloid cells, B/plasma cells, and tissue cells. **j**, Spatial distribution of predicted CXCL9–CXCR3 ligand-receptor. **k**, Zoom-in of a representative region showing identified niche of T cells and macrophages. Cells are colored according to their cluster assignments. **l**, Violin plots showing the expression levels of T- and macrophage-related genes across different cell clusters. **m**, Spatial visualization of the expression levels of T- and macrophage-related genes at the matched histopathological locations. Color scale indicates normalized expression levels.

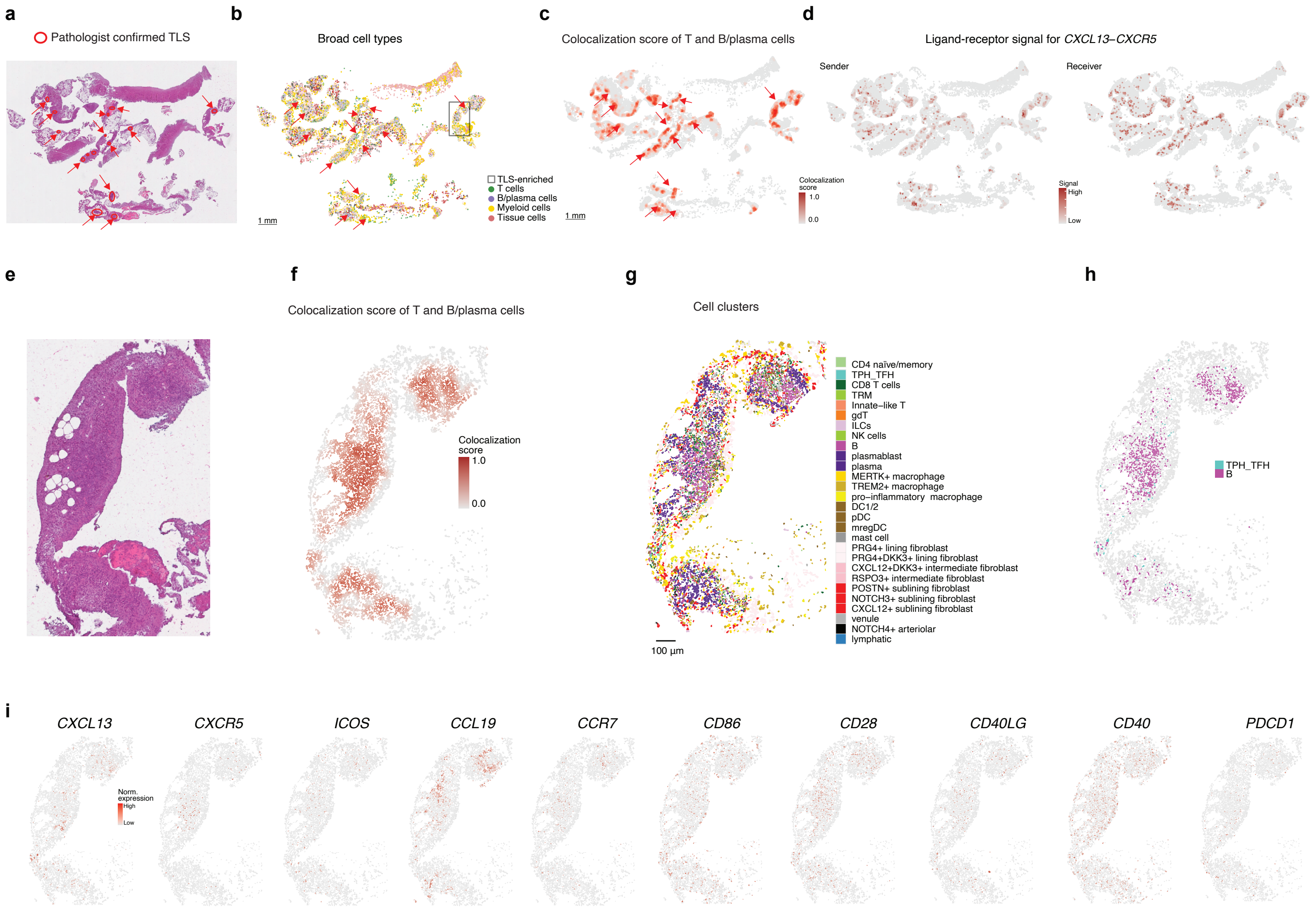
a
Pathologist confirmed TLS
b
Broad cell types
TLS-enriched
T cells
B/plasma cells
Myeloid cells
Tissue cells
1 mm
c
Colocalization score of T and B/plasma cells
Colocalization score
1.0
0.0
1 mm
d
Ligand-receptor signal for CXCL13–CXCR5
Sender
Receiver
Signal
High
Low
e
f
Colocalization score of T and B/plasma cells
Colocalization score
1.0
0.0
g
Cell clusters
CD4 naïve/memory
TPH_TFH
CD8 T cells
TRM
Innate–like T
gdT
ILCs
NK cells
B
plasmablast
plasma
MERTK+ macrophage
TREM2+ macrophage
pro–inflammatory macrophage
DC1/2
pDC
mregDC
mast cell
PRG4+ lining fibroblast
PRG4+DKK3+ lining fibroblast
CXCL12+DKK3+ intermediate fibroblast
RSPO3+ intermediate fibroblast
POSTN+ sublining fibroblast
NOTCH3+ sublining fibroblast
CXCL12+ sublining fibroblast
venule
NOTCH4+ arteriolar
lymphatic
100 µm
h
TPH_TFH
B
i
CXCL13
CXCR5
ICOS
CCL19
CCR7
CD86
CD28
CD40LG
CD40
PDCD1
Norm. expression
High
Low

**Supplemental Figure 5: Identified TLS-clusters in JIA synovium. a**, H&E staining images of synovial tissue, illustrating representative TLS as identified by a blinded pathologist. **b**, Spatial distribution of T and B cells. Scale bar, 1 mm. The highlighted black square indicates the TLS-enriched region. **c**, Colocalization scores for T and B cells. **d**, Spatial *CXCL13-CXCR5* interaction signals. **e**, H&E staining image of zoomed-in of the area with the highest TLS density highlighted in panel **b. f-h**, Same region as in panel **e**, colored by T and B cell colocalization score (**f**) and fine-scale cell cluster assignments (**g-h**). **i**, Spatial expression patterns of TLS-associated genes.

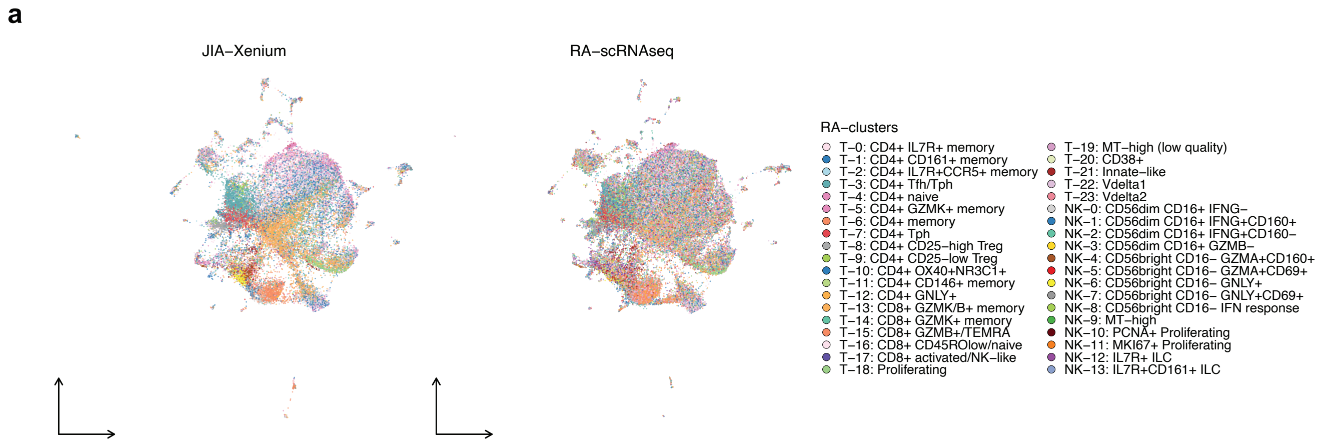
a
JIA–Xenium
RA–scRNAseq
RA–clusters
T–0: CD4+ IL7R+ memory
T–1: CD4+ CD161+ memory
T–2: CD4+ IL7R+CCR5+ memory
T–3: CD4+ Tfh/Tph
T–4: CD4+ naive
T–5: CD4+ GZMK+ memory
T–6: CD4+ memory
T–7: CD4+ Tph
T–8: CD4+ CD25–high Treg
T–9: CD4+ CD25–low Treg
T–10: CD4+ OX40+NR3C1+
T–11: CD4+ CD146+ memory
T–12: CD4+ GNLY+
T–13: CD8+ GZMK/B+ memory
T–14: CD8+ GZMK+ memory
T–15: CD8+ GZMB+/TEMRA
T–16: CD8+ CD45ROlow/naive
T–17: CD8+ activated/NK–like
T–18: Proliferating
T–19: MT–high (low quality)
T–20: CD38+
T–21: Innate–like
T–22: Vdelta1
T–23: Vdelta2
NK–0: CD56dim CD16+ IFNG–
NK–1: CD56dim CD16+ IFNG+CD160+
NK–2: CD56dim CD16+ IFNG+CD160–
NK–3: CD56dim CD16+ GZMB–
NK–4: CD56bright CD16– GZMA+CD160+
NK–5: CD56bright CD16– GZMA+CD69+
NK–6: CD56bright CD16– GNLY+
NK–7: CD56bright CD16– GNLY+CD69+
NK–8: CD56bright CD16– IFN response
NK–9: MT–high
NK–10: PCNA+ Proliferating
NK–11: MKI67+ Proliferating
NK–12: IL7R+ ILC
NK–13: IL7R+CD161+ ILC

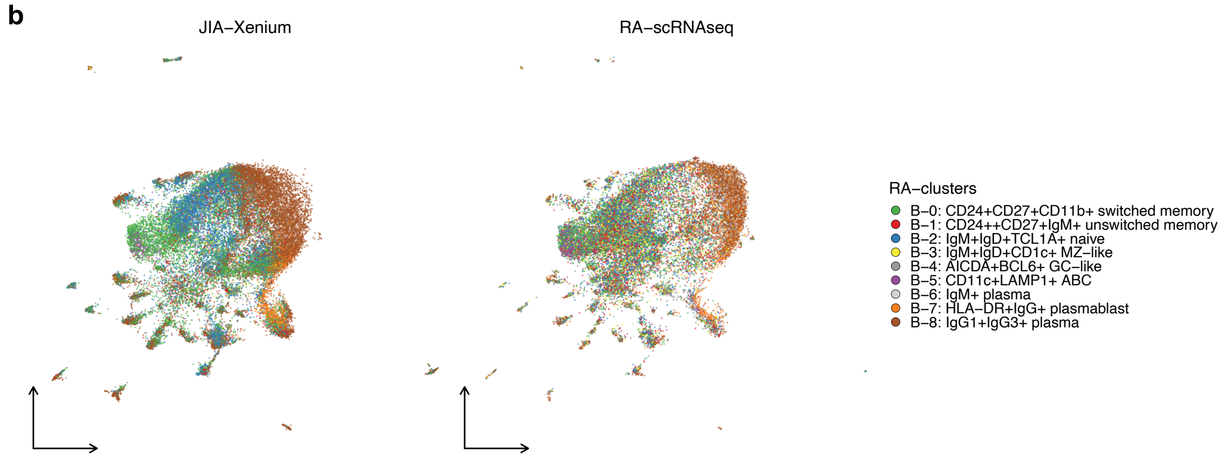
b
JIA–Xenium
RA–scRNAseq
RA–clusters
B–0: CD24+CD27+CD11b+ switched memory
B–1: CD24++CD27+IgM+ unswitched memory
B–2: IgM+IgD+TCL1A+ naive
B–3: IgM+IgD+CD1c+ MZ–like
B–4: AICDA+BCL6+ GC–like
B–5: CD11c+LAMP1+ ABC
B–6: IgM+ plasma
B–7: HLA–DR+IgG+ plasmablast
B–8: IgG1+IgG3+ plasma

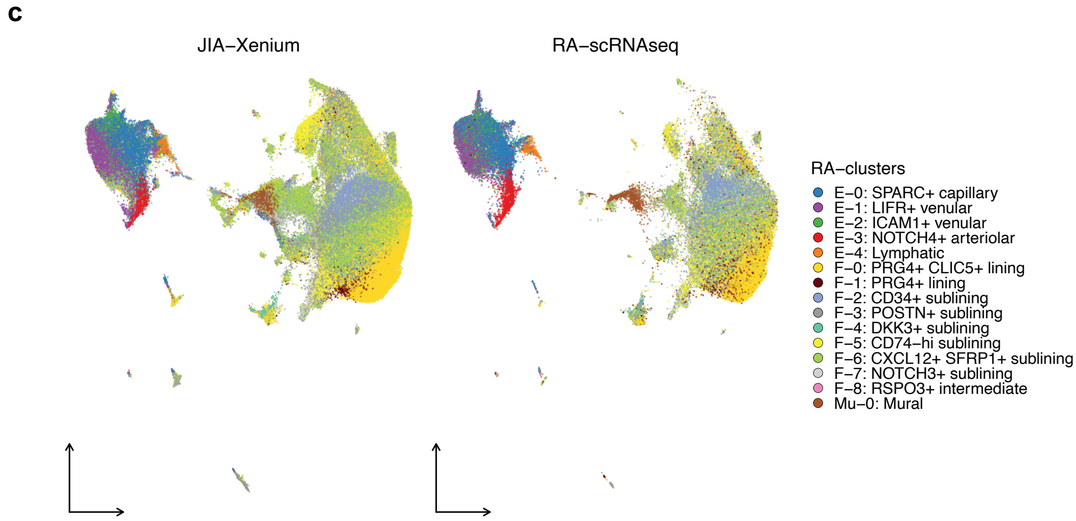
c
JIA–Xenium
RA–scRNAseq
RA–clusters
E–0: SPARC+ capillary
E–1: LIFR+ venular
E–2: ICAM1+ venular
E–3: NOTCH4+ arteriolar
E–4: Lymphatic
F–0: PRG4+ CLIC5+ lining
F–1: PRG4+ lining
F–2: CD34+ sublining
F–3: POSTN+ sublining
F–4: DKK3+ sublining
F–5: CD74–hi sublining
F–6: CXCL12+ SFRP1+ sublining
F–7: NOTCH3+ sublining
F–8: RSPO3+ intermediate
Mu–0: Mural

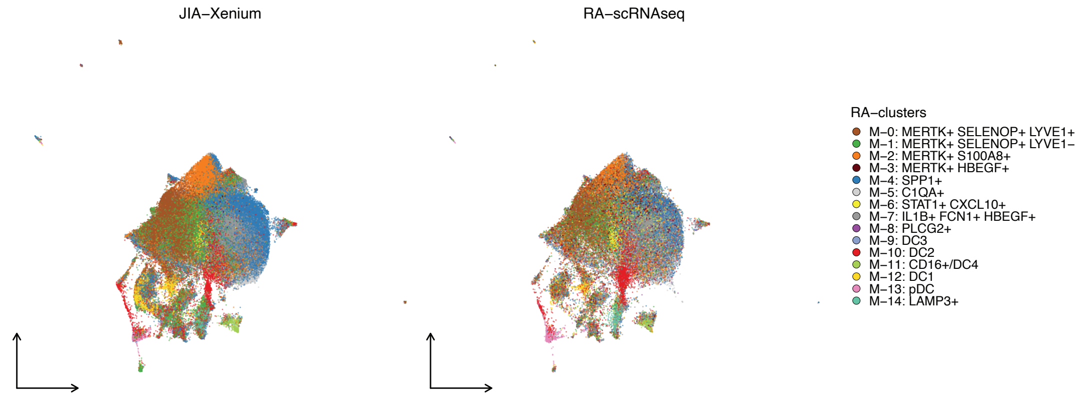
JIA–Xenium
RA–scRNAseq
RA–clusters
M–0: MERTK+ SELENOP+ LYVE1+
M–1: MERTK+ SELENOP+ LYVE1–
M–2: MERTK+ S100A8+
M–3: MERTK+ HBEGF+
M–4: SPP1+
M–5: C1QA+
M–6: STAT1+ CXCL10+
M–7: IL1B+ FCN1+ HBEGF+
M–8: PLCG2+
M–9: DC3
M–10: DC2
M–11: CD16+/DC4
M–12: DC1
M–13: pDC
M–14: LAMP3+

**Supplemental Figure 6: Cell type mapping between JIA Xenium spatial transcriptomics and RA synovial scRNA-seq data across immune and stromal compartments. a-d**: UMAP projections of JIA synovium (Xenium, left) and RA synovium (scRNA-seq, right) showing StabMap-based label transfer for major immune and stromal compartments. Each cell is colored according to the RA scRNA-seq-derived reference cluster annotation transferred to Xenium cells using k-nearest neighbor (kNN) mapping in a shared latent space. **a**: T/ILCs, **b**: B cell and plasma cell subsets, **c**: Fibroblast and endothelial cell subsets, and **d**: Myeloid cell subsets.
