## Supplemental Table 1 for "Subcellular spatial transcriptomics reveals immune–stromal crosstalk within the synovium of patients with juvenile idiopathic arthritis"

**Supplementary Table 1: Clinical features of the patients**

| Patient | JIA 1 | JIA 2 | JIA 3 | JIA 4 | JIA 5 | JIA 6 | JIA 7 | JIA 8 | JIA 9 |
|---|---|---|---|---|---|---|---|---|---|
| **Sex** | F | F | M | F | M | F | F | F | F |
| **Disease Duration (years)** | 0.5 | 6 | 3 | 0.2 | 0.5 | 4 | 0.2 | 16 | 0.5 |
| **Time from flare onset to biopsy (years)** | 0.1 | 0.1 | 0.3 | 0.2 | 0.5 | 0.2 | 0.2 | 0.1 | 0.5 |
| **JIA type** | Oligo | Oligo | Poly | Oligo | Poly | Poly | Oligo | Oligo | Oligo |
| **Source of Synovium** | Knee | Knee | Knee | Knee | Knee | Knee | Knee | Knee | Knee |
| **Prior treatment** | Steroid injection | Steroid injection | MTX, LEF, ADA | None | None | Steroid injection, MTX | None | MTX, ETA, ADA | MTX |
| **Systemic treatment at the time of biopsy** | None | MTX | LEF, ADA | None | None | MTX | None | none | MTX |
| **Uveitis** | Negative | Negative | Positive | Negative | Negative | Negative | Negative | Positive | Negative |
| **ANA** | Positive | Positive | Positive | Positive | Positive | Positive | Positive | Negative | Positive |
| **RF** | Negative | Negative | Negative | Negative | Negative | Negative | Negative | Negative | Negative |
| **CRP (mg/L)** | 0 | 35 | 29 | 11 | 0 | 0 | 0 | 0 | 0 |
| **Krenn score (total)** | 6 | 2 | 5 | 8 | 7 | 6 | 8 | 4 | 6 |
| **Krenn lining** | 2 | 0 | 1 | 2 | 2 | 2 | 3 | 1 | 2 |
| **Krenn inflammation** | 2 | 1 | 2 | 3 | 3 | 2 | 3 | 1 | 2 |
| **Krenn stroma** | 2 | 1 | 2 | 3 | 2 | 2 | 2 | 2 | 2 |

Oligo: oligoarticular juvenile idiopathic arthritis, Poly: polyarticular juvenile idiopathic arthritis, MTX: methotrexate, LEF: leflunomide, ETA: etanercept, ADA: adalimumab, ANA: antinuclear antibody, RF: rheumatoid factor, CRP: C-reactive protein
